# Durability analysis of the highly effective BNT162b2 vaccine against COVID-19

**DOI:** 10.1101/2021.09.04.21263115

**Authors:** Arjun Puranik, Patrick J. Lenehan, John C. O’Horo, Michiel J.M. Niesen, Abinash Virk, Melanie D. Swift, Walter Kremers, A.J. Venkatakrishnan, Joel E. Gordon, Holly L. Geyer, Leigh Lewis Speicher, Venky Soundararajan, Andrew D. Badley

## Abstract

SARS-CoV-2 breakthrough infections have been increasingly reported in fully vaccinated individuals. We conducted a test-negative case-control study to assess the durability of protection after full vaccination with BNT162b2, defined as 14 days after the second dose, against polymerase chain reaction (PCR)-confirmed symptomatic SARS-CoV-2 infection, in a national medical practice between February 1, 2021 and August 22, 2021. We fit conditional logistic regression (CLR) models stratified on residential county and calendar time of testing to assess the association between time elapsed since vaccination and the odds of symptomatic infection or non-COVID-19 hospitalization (negative control), adjusted for several covariates. The primary population included 652 individuals who had a positive symptomatic test after full vaccination with BNT162b2 (cases) and 5,946 individuals with at least one negative symptomatic test after full vaccination (controls). The adjusted odds of symptomatic infection were higher 120 days after full vaccination versus at the date of full vaccination (Odds Ratio [OR]: 3.21, 95% confidence interval [CI]: 1.33-7.74). Importantly, the odds of infection were still lower 150 days after the first BNT162b2 dose as compared to 4 days after the first dose (OR: 0.3, 95% CI: 0.19-0.45), when immune protection approximates the unvaccinated status. Low rates of COVID-19 associated hospitalization or death in this cohort precluded analyses of these severe outcomes. The odds of experiencing a non-COVID-19 hospitalization decreased with time since vaccination, suggesting a possible underestimation of waning protection by this approach due to confounding factors. Taken together, these data constitute an early signal for waning protection against symptomatic illness while also providing reassurance that BNT162b2 continues to protect against symptomatic SARS-CoV-2 infection several months after full vaccination. Continued surveillance of COVID-19 vaccine durability, particularly against severe disease, is critical to guide effective and equitable strategies to respond to the pandemic, including distribution of booster doses, development of new vaccines, and implementation of both pharmaceutical and nonpharmaceutical interventions.

## Introduction

Infection with severe acute respiratory syndrome coronavirus 2 (SARS-CoV-2) and the resulting coronavirus disease-2019 (COVID-19) have impacted over 215 million people worldwide, resulting in more than 4.5 million deaths to date.^1^ Efforts were rapidly initiated during the early months of the pandemic to develop vaccines that would reduce community transmission and disease burden, leading to the clinical testing and subsequent Emergency Use Authorization (EUA) by the United States Food and Drug Administration (FDA) of three vaccines by February 2021: BNT162b2 (mRNA vaccine by Pfizer-BioNTech authorized in December 2020), mRNA-1273 (mRNA vaccine by Moderna authorized in December 2020), and Ad26.COV2.S (adenoviral vector vaccine by Janssen authorized in February 2021).^2–6^ As of September 3, 2021, over 445 million vaccine doses have been administered to over 205 million people in the United States, with over 60% of the adult population reaching full vaccination status per the Centers for Disease Control and Prevention (CDC) definition.^7^

Randomized phase 3 clinical trials demonstrated over 90% efficacy in preventing symptomatic infection for both mRNA vaccines and approximately 65% efficacy in the same for Ad26.COV2.S.^3,5,6^ Subsequent analyses of individuals vaccinated outside the trial setting have yielded similar results, and BNT162b2 was recently granted full approval by the FDA for individuals 16 years of age and older.^8–11^ However, especially with the continued evolution of new SARS-CoV-2 strains, including the Delta variant which is now the dominant strain in the United States and worldwide, it is important to keep monitoring vaccine effectiveness over time and against specific variants. Indeed, several studies have suggested declining protection against infection during recent months.^12–19^ Although this change could be related to several factors in addition to waning immune protection such as changes in non-pharmaceutical interventions (e.g., masking, social distancing, travel restrictions) or more efficient immune evasion by the Delta variant, these early signals have prompted the recommendation of mRNA vaccine booster doses for immunocompromised individuals in the United States and elsewhere.^20,21^ Further understanding of COVID-19 vaccine durability will inform strategies regarding whether and when certain individuals should consider receiving a booster dose. Here we use test-negative case-control designs to assess the durability of BNT162b2 among individuals who were vaccinated and subsequently tested for suspected SARS-CoV-2 infection at the Mayo Clinic.

## Methods

### Study population

This is a retrospective study of individuals who were vaccinated with BNT162b2 between February 1, 2021 and August 22, 2021, and who subsequently underwent polymerase chain reaction (PCR) testing for suspected symptomatic SARS-CoV-2 infection at the Mayo Clinic. According to the Centers for Disease Control and Prevention (CDC), full vaccination with BNT162b2 is defined as beginning 14 days after the second dose.^22^ This study was reviewed and deemed exempt by the Mayo Clinic institutional review board. Those who had specifically opted out of inclusion of electronic medical records in research were excluded. Inclusion and exclusion criteria were defined as follows.

#### Inclusion criteria

1. Age greater than or equal to 18 years as of February 1, 2021.
2. Received two doses of BNT162b2 per-protocol, with the first dose administered on or after February 1, 2021. Per-protocol BNT162b2 vaccination was defined as two doses administered 18-28 days apart with no doses of other COVID-19 vaccines (i.e. mRNA-1273, Ad26.COV2.S) administered at any time before the second dose.
3. At least one clinical encounter at the Mayo Clinic in the three years preceding the study start date (i.e. between February 1, 2018 and February 1, 2021), per the electronic health record.

#### Exclusion criteria

1. Any positive SARS-CoV-2 PCR test prior to the date of full vaccination.

The derivation of this study population is illustrated in **Figure 1** and **Figure S1**, and the demographic and clinical characteristics of the cohort along with the underlying fully vaccinated population is shown in **Table 1**.

**Table 1.**
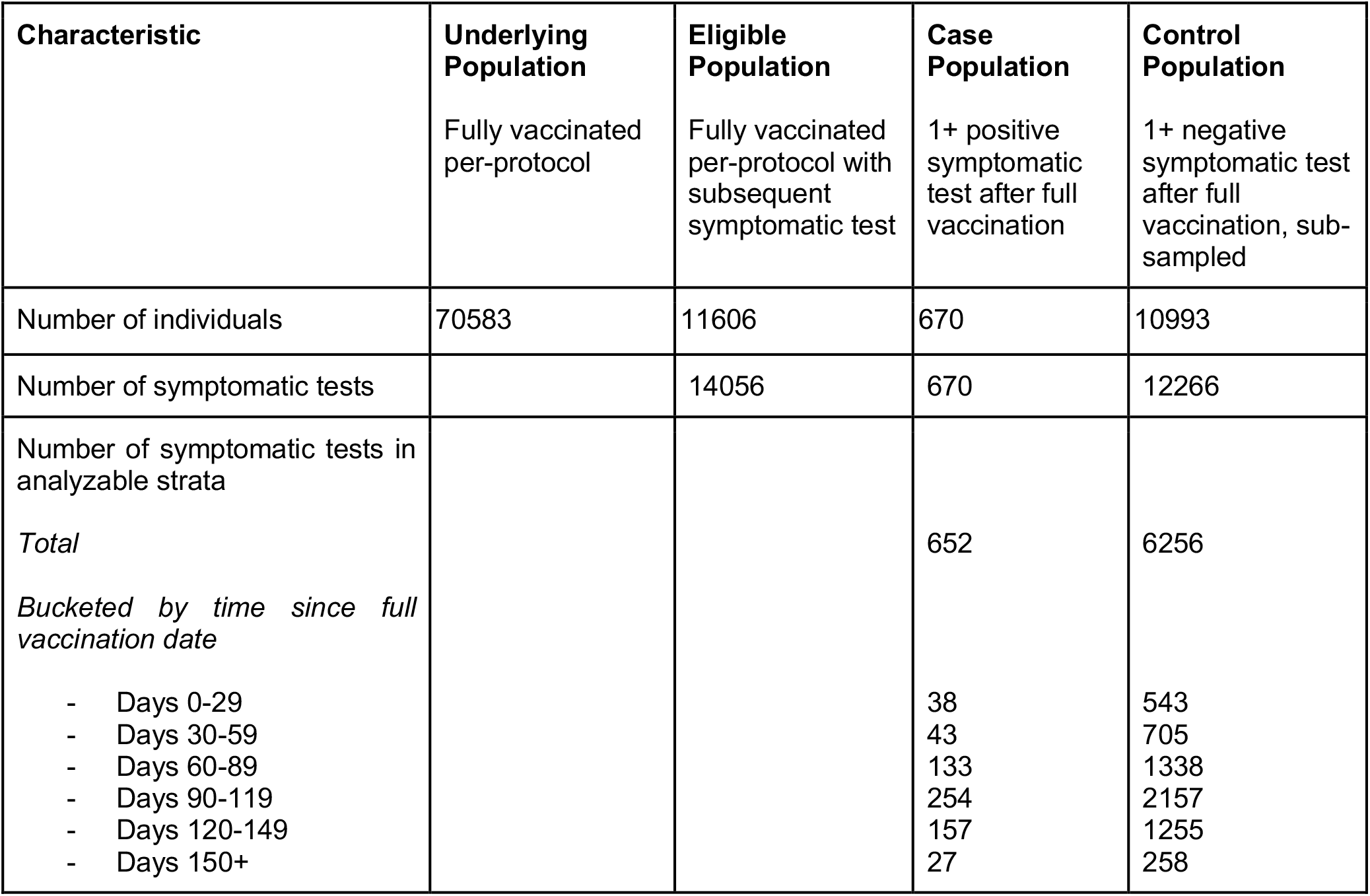
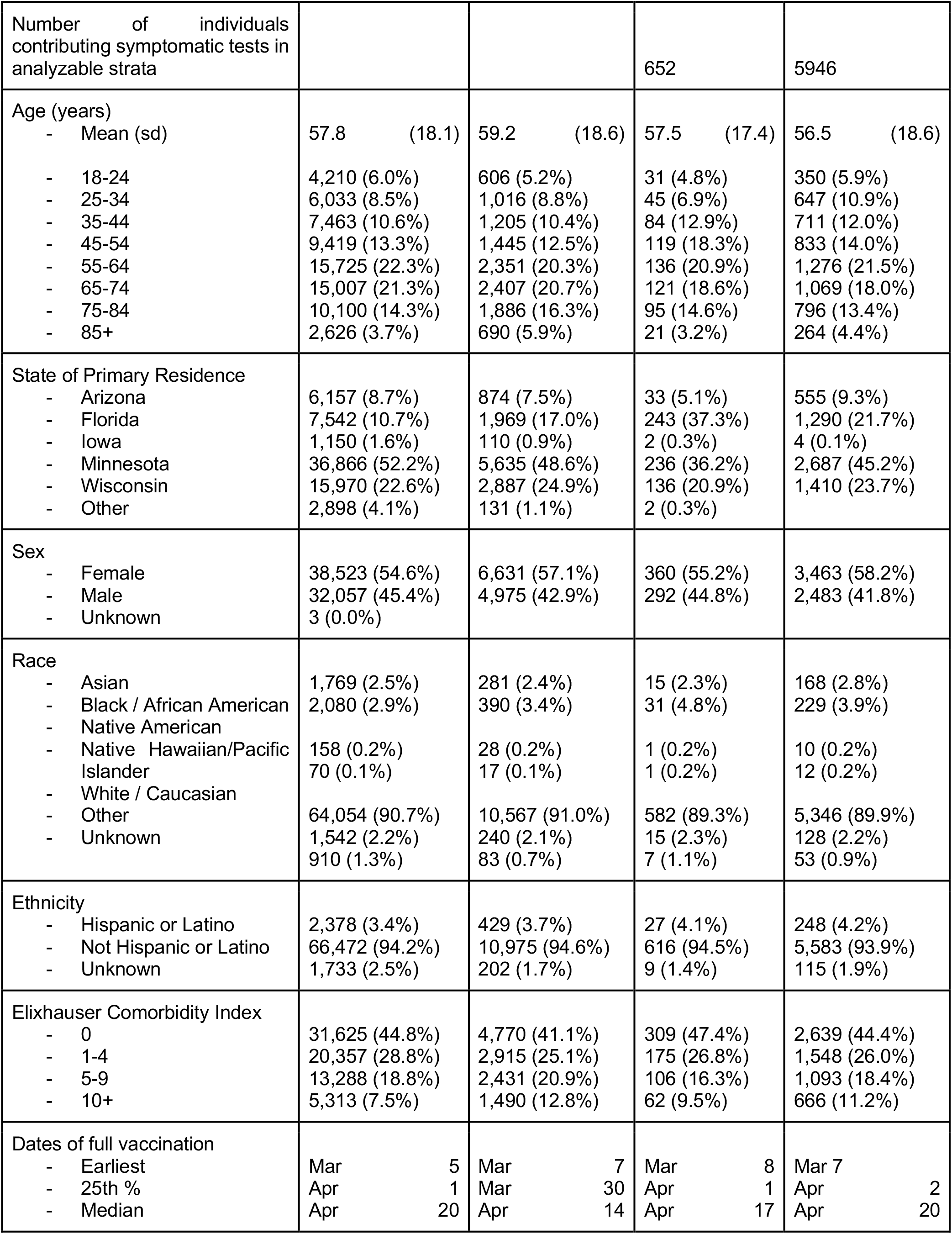
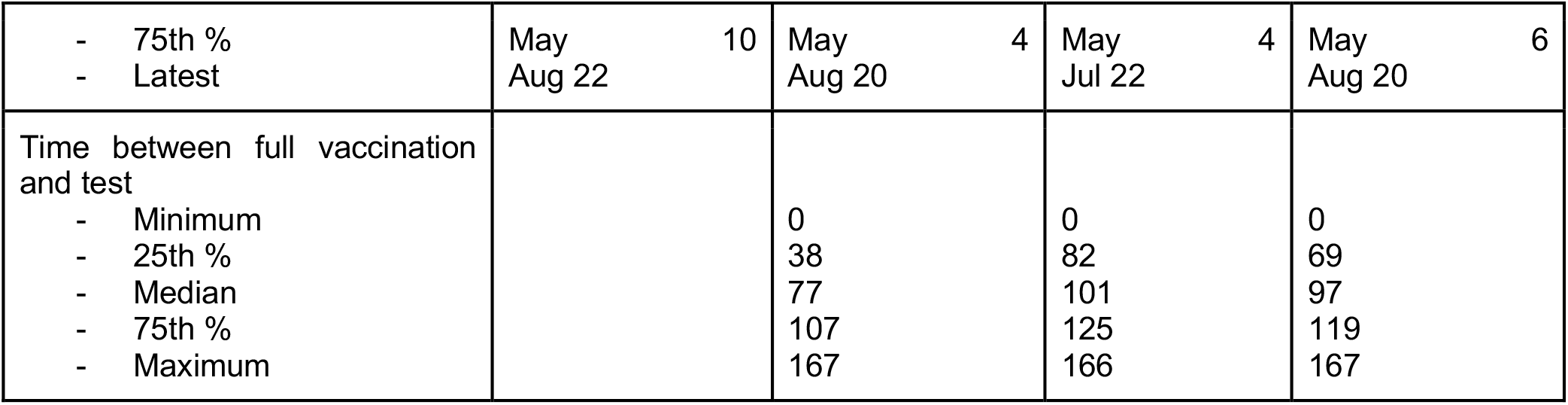
Demographic and clinical characteristics of cases and controls for primary analysis of symptomatic infection among fully vaccinated BNT162b2 recipients. The underlying population corresponds to the set of individuals who received their first BNT162b2 dose on or after February 1, 2021 and were fully vaccinated per protocol (i.e. with two doses administered 18-28 days apart and with no prior positive SARS-CoV-2 PCR tests before the date of full vaccination). The eligible population corresponds to the subset of the underlying population who underwent at least one symptomatic test after the date of full vaccination. Cases correspond to the first positive symptomatic test for a given individual in the eligible population; by definition, the number of individuals contributing cases is the same as the number of cases. Controls correspond to negative symptomatic tests after full vaccination which occur before the given individual has experienced any positive SARS-CoV-2 PCR tests; an individual can contribute multiple controls during the study period, so the number of individuals in the control population is less than the total number of tests (controls) contributed. Because an individual can contribute negative tests (controls) prior to contributing a positive test, the number of individuals in the eligible population is smaller than the sum of the number of individuals in the case and control populations. Sub-sampling in the control population refers to the process in which negative tests from a given individual were (i) excluded if they occurred after a positive test or within the 15 days before a positive test (possible false negative), (ii) randomly sampled if they occurred within 15 days of each other (possibly during the same symptomatic illness), and (iii) randomly sampled if the individual contributed more than three negative tests during the study period. A stratum (defined by the regression equation as a unique combination of county and calendar week of testing) is considered analyzable if it includes at least one case and at least one control, because strata including only cases or only controls do not contribute to the estimation of the regression coefficients. For all cases and controls, all summarized characteristics correspond to only individuals who contributed at least one symptomatic test to an analyzable stratum.

**Figure 1.**
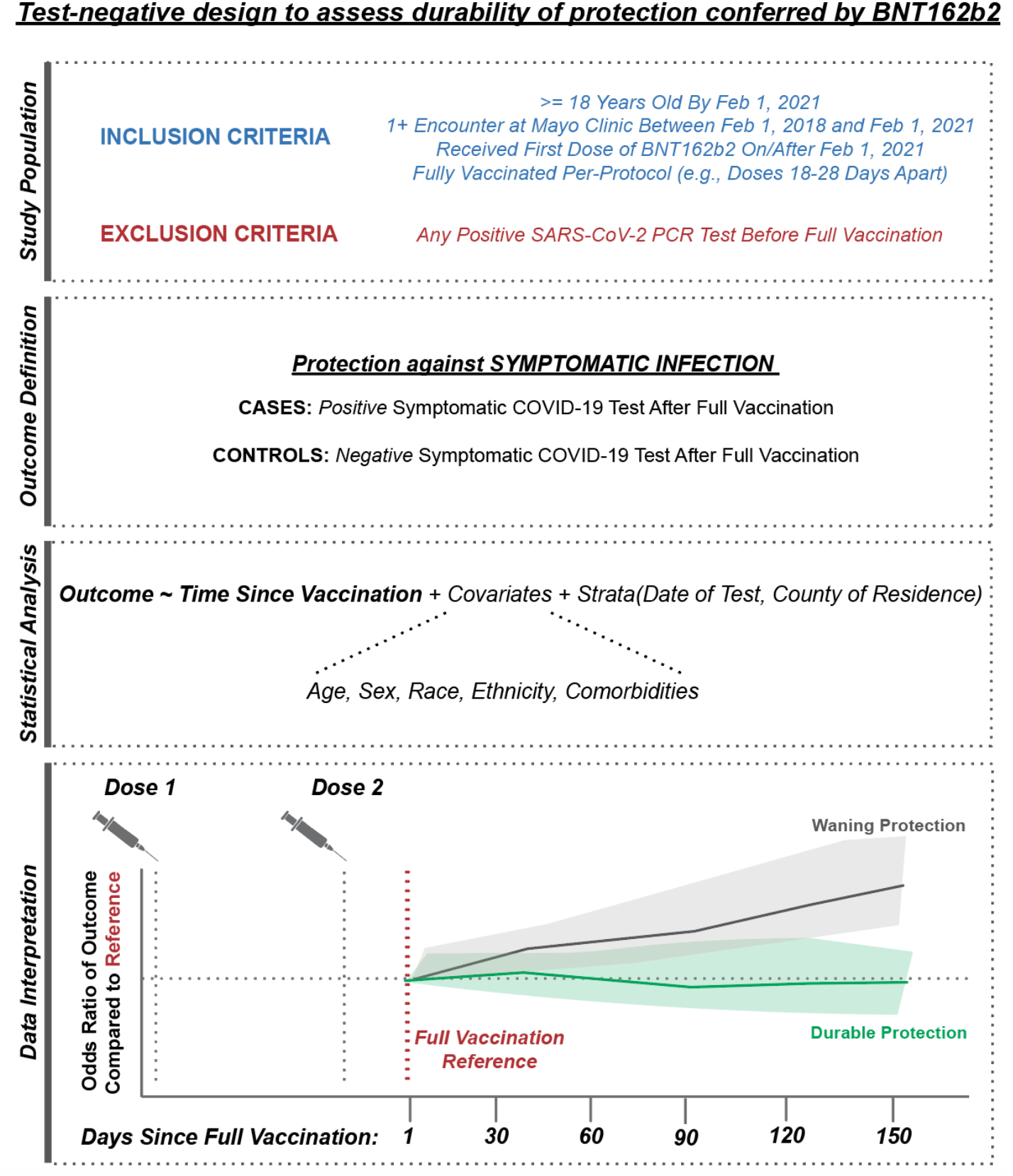
Schematic representation of study design. From top to bottom, (i) inclusion and exclusion criteria to define the population eligible for this test-negative analysis, (ii) definition of the clinical outcomes of interest, (iii) framework for statistical analysis, and (iv) schematic representation of data interpretation. (i) Individuals were included if they were at least 18 years old, had a record of at least one encounter at the Mayo Clinic in the three years prior to the study start date, and were fully vaccinated per-protocol with BNT162b2 (with the first dose administered on or after February 1, 2021), and underwent at least one symptomatic SARS-CoV-2 PCR test after the date of full vaccination. Individuals were excluded if they received a positive PCR test prior to their date of full vaccination. (ii) The outcome was defined as symptomatic SARS-CoV-2 infection, and cases and controls were defined accordingly. (iii) Conditional logistic regression was used to assess the potential relationship between the odds of experiencing symptomatic infection and time since vaccination, while accounting for other clinical and demographic covariates. (iv) The odds of symptomatic infection were assessed over time after full vaccination (modeled as a linear spline) relative to the odds at the date of full vaccination, which is expected to correspond to maximal vaccine-mediated protection. An increase in the odds ratio with time since vaccination would be interpreted as evidence for waning protection, while a consistent odds ratio over (relative) time would be interpreted as durable protection.

### Study design

We performed a test-negative case-control analysis to assess whether the protection conferred by BNT162b2 wanes over time, similar to a study design described previously to analyze intraseason waning effectiveness of influenza vaccination.^23^ To do so, we used conditional logistic regression to assess the odds of symptomatic SARS-CoV-2 infection and one negative control outcome (non-COVID-19 hospitalization) over time after full vaccination, while adjusting for relevant covariates. Symptomatic infection was defined as a positive result from a SARS-CoV-2 PCR test that was not designated as “asymptomatic” by the ordering provider (subsequently referred to as “symptomatic tests”).^22^ Because we expect the date of full vaccination to approximate the time of maximal protection, we assess vaccine durability by estimating the odds of symptomatic infection at 30, 60, 90, 120, and 150 days after this time point.

### Definitions of cases, controls, and at-risk time

Cases were defined as the first positive symptomatic test for a given individual; if an individual contributed multiple positive tests, only their first test was included as a case. Controls were defined as negative symptomatic tests which did not occur after any prior positive SARS-CoV-2 PCR tests (asymptomatic or symptomatic). Individuals who met the inclusion and exclusion criteria outlined above were eligible to contribute cases and controls from their date of full vaccination until they (i) had any positive test result (symptomatic or asymptomatic), (ii) received a third dose of any COVID-19 vaccine (BNT162b2, mRNA-1273, or Ad26.COV2.S), (iii) died, or (iv) reached the end of the study period. If an individual contributed a negative symptomatic test 15 or fewer days before a positive test, that negative test was excluded as a possible false negative. If an individual contributed multiple negative symptomatic tests within 15 days of each other, then one of those tests was randomly selected as a control while the others were dropped; this step was taken to avoid counting multiple controls from a potential single symptomatic illness. Further, if an individual contributed more than three negative symptomatic tests over the study duration, then three tests were randomly selected as controls while the others were dropped, as was recently described in a test-negative case-control study of COVID-19 vaccine effectiveness.^24^

As a negative control analysis, we assessed protection against non-COVID-19 hospitalization, an outcome which we do not expect to be impacted by time since vaccination. In other test negative designs on influenza, other respiratory infections were used as the negative control.^23^ Because of the myriad impacts of the pandemic and nonpharmaceutical interventions on other respiratory infections, such an approach may not be valid here.^25^ Although non-COVID-19 related hospitalization could be impacted by factors such as changes in healthcare-seeking behavior (including elective procedures) after vaccination, it appeared to be a reasonable negative control to evaluate. Here, cases were defined as instances in which an individual experienced a negative symptomatic test (i.e. ruled out for COVID-19 diagnosis) and was subsequently admitted to the hospital within 14 days. Controls were defined as instances in which an individual experienced a negative symptomatic test and was not subsequently admitted to the hospital within 14 days. Individuals who met the inclusion and exclusion criteria outlined above were eligible to contribute cases and controls from their date of full vaccination until they (i) were hospitalized within 14 days of a negative symptomatic test, (ii) received a third dose of any COVID-19 vaccine (BNT162b2, mRNA-1273, or Ad26.COV2.S), (iii) died, or (iv) reached the end of the study period. The same rules were applied as described above for cases in which an individual contributed (i) a negative test shortly before contributing a positive test, (ii) multiple negative symptomatic tests within 15 days of each other, or (iii) more than three negative symptomatic tests over the duration of the study. Because 14 days of follow-up were required after a positive symptomatic test to observe this outcome, cases and controls were only considered from tests that were performed on or before August 8, 2021 (14 days before the last date of data collection).

### Primary exposure, covariates, and stratification factors

Variables that are potentially associated with the likelihood of eligibility for vaccination at a given time, seeking out vaccination, testing positive for SARS-CoV-2, or experiencing severe COVID-19 were included as covariates or stratification variables in the regression models. The primary exposure of interest and each such other variable, denoted as X_1_-X_8_ in the regression equation listed in the *Statistical Analysis* section, is described below.

#### Primary exposure

1. X_1_: Time since full vaccination, defined as the number of days between the symptomatic PCR test and the date of full vaccination. This variable was modeled as a linear spline with knots at 30, 60, 90, 120, and 150 days since full vaccination. As described above, the date of full vaccination is expected to correspond to a time of maximal protection and thus was considered as the reference. Results are presented as the odds of symptomatic infection at each knot relative to this reference.

#### Covariates

2. X_2_: Age in years as of the study start date (February 1, 2021), modeled as a linear spline with knots at 25, 35, 45, 55, 65, 75, and 85 years. The minimum age (18 years old) was considered the reference. Results are presented as the odds of symptomatic infection at each subsequent knot relative to this reference.
3. X_3_: Elixhauser Comorbidity Index score (“Elixhauser score”), categorized into four groups (0, 1-4, 5-9, and 10+). A score of 0 was considered the reference category.
4. X_4_: Race, categorized into seven groups (listed alphabetically: Asian, Black/African American, Native American, Native Hawaiian/Pacific Islander, Other, White/Caucasian, Unknown). Whie/Caucasian was considered the reference category because it comprised the majority of individuals in the study.
5. X_5_: Ethnicity, categorized into three groups (listed alphabetically: Hispanic/Latino, not Hispanic/Latino, Unknown). “Not Hispanic/Latino” was considered the reference category because it comprised the majority of individuals in the study.
6. X_6_: Sex, categorized into three groups (listed alphabetically: female, male, and unknown). Female was considered the reference category.

#### Stratification factors

7. X_7_: County of residence at the time of testing for the individual who underwent the symptomatic test.
8. X_8_: Calendar time of test, categorized in one-week intervals starting on the date of the first symptomatic test after full vaccination (here, starting on March 7, 2021).

### Determination of Elixhauser Comorbidity Index score

We used the *comorbidity* package (version 0.5.3) in R (version 4.1.0, www.r-project.org, Vienna, Austria) to identify ICD-9 and ICD-10 codes that correspond to each Elixhauser comorbidity. For each individual, we extracted all such diagnosis codes in the Mayo Clinic electronic health record from the five years preceding this study (i.e. between February 1, 2016 and February 1, 2021). The Elixhauser score was defined as the total number of Elixhauser comorbidities present in at least one record during this five year period. For subsequent analyses, these values were bucketed as described above (0, 1-4, 5-9, ≥10).

### Statistical analysis

Briefly, for each outcome (i.e. symptomatic SARS-CoV-2 infection and non-COVID-19 hospitalization), we fit a conditional logistic regression (CLR) model to estimate the odds of experiencing the outcome of interest each day after the date of full vaccination compared to the odds of experiencing that outcome on the date of full vaccination, while adjusting for the covariates described above.

The CLR models were each defined by the equation, 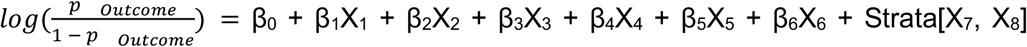, where the covariates and conditioning variables X_1_-X_8_ are described in the section above.

Models were fit using the *clogit* function from the *survival* package (version 3.2.11) in R (Version, 4.1.0, www.r-project.org, Vienna, Austria). Confidence intervals and tests were based upon the Wald method. Odds ratios were considered statistically significant if the confidence interval did not include 1.

### Secondary analysis: odds of infection relative to the first vaccine dose rather than full vaccination

The primary analysis estimates the change in odds of infection over time relative to the time of maximal vaccine protection without providing the important context of the baseline risk of infection in the absence of vaccination. In our study population, which consists entirely of vaccinated individuals with no prior history of COVID-19 diagnosis, it is not possible to directly measure this risk of infection in the unvaccinated state. However, the protective effect of BNT162b2 sets in after approximately two weeks, and we reasoned that the risk of infection in the unvaccinated state could be approximated by the risk of infection shortly after initial vaccination (e.g., within days 4-10 after the first dose).^3,9^ We thus modified the inclusion and exclusion criteria criteria from our primary analysis as follows.

#### Inclusion criteria

1. Age greater than or equal to 18 years as of February 1, 2021.
2. Received at least one dose of BNT162b2 on or after February 1, 2021.
3. At least one clinical encounter at the Mayo Clinic in the three years preceding the study start date (i.e. between February 1, 2018 and February 1, 2021), per the electronic health record.

#### Exclusion criteria

1. Any positive SARS-CoV-2 PCR test prior to the date of full vaccination.
2. Received one or more doses of another COVID-19 vaccine (mRNA-1273 or Ad26.COV2.S) on or before February 1, 2021.

Cases and controls were defined as described above but with modifications to the censoring protocol. Specifically, individuals who met the inclusion and exclusion criteria were eligible to contribute cases and controls from four days after their first vaccine dose until they (i) had any positive test result (symptomatic or asymptomatic), (ii) went off-protocol for their vaccination regimen (i.e. received a second dose of BNT162b2 less than 18 days after the first dose, did not receive a second dose of BNT162b2 by 28 days after their first dose, or received a dose of a different COVID-19 vaccine within 28 days of their first dose), (iii) received a third dose of any COVID-19 vaccine, (iv) died, or (v) reached the end of the study period. Note that at-risk time was defined to begin four days after the first dose (rather than the first day after the first dose) for two reasons: (i) individuals with respiratory symptoms were often encouraged to delay vaccination, resulting in a potential bias toward lower symptomatic infection rates immediately following vaccination; and (ii) individuals who develop symptomatic COVID-19 shortly after vaccination may attribute their symptoms to vaccine side effects, resulting in a likely delay of testing.

The same CLR model described above for the main analysis was applied, except that the “Time since vaccination” variable was now modeled as a linear spline with eight knots: 10, 15, 21, 35, 60, 90, 120, 150, and 180 days after the first dose. It is recommended that the second dose of BNT162b2 is administered 21 days after the first, with full vaccination thus expected to start 35 days after the first dose. Because the vaccine is not expected to provide protection until about two weeks after the first dose, we considered 4 days after the first dose as a reference time point to approximate unvaccinated status.^3,5,9^ Results are presented as the odds of symptomatic infection at each knot relative to this reference.

### Sensitivity analyses

We performed two sensitivity analyses to assess the robustness of the results derived from the primary and secondary analyses.

#### 1. Age subgroup analysis relative to the first vaccine dose

While the previous approaches included age as a covariate, we considered it important to determine whether any observed signal of waning was observed across all age groups. We thus divided the cohort of individuals vaccinated with one or two doses of BNT162b2 (i.e. the cohort from the secondary analysis) into three age groups: 18-44, 45-64, and ≥65 years. For each age group, we then fit the same CLR model as was described above for the secondary analysis (still including age as a covariate) to evaluate the association between time since first vaccination (i.e. four days after the first dose) and the odds of symptomatic infection or non-COVID-19 hospitalization.

#### 2. Stratification by time of vaccination rather than by time of testing

By performing CLR with stratification by the time of testing, individuals with more time since vaccination in any given stratum will inherently have been vaccinated at earlier times during the vaccine rollout. Because the time of vaccination is likely associated with the risk of SARS-CoV-2 infection and the likelihood of engagement with the healthcare system, this approach could yield biased estimates of the infection odds over time after vaccination. We thus performed a sensitivity analysis in which the CLR was stratified by county, date of full vaccination (in two-week calendar intervals), and county-level COVID-19 incidence at the time of testing, rather than on county and time of testing (as was the case in the previous model). Here, we also included an additional covariate to capture the dominant SARS-CoV-2 variant at the time of the test; this was not included in our primary model because the variant prevalence was implicitly captured by stratifying on the time of testing. Specifically, the additional variables considered here were:

##### Covariates

1. X_9_: Dominant SARS-CoV-2 variant, categorized as Alpha, Delta, Neither, or Unknown. The prevalence of Alpha and Delta variants was determined for each state in twice-monthly intervals (i.e. from the first to the 15th day of each month, and from the 16th to the last day of each month) using publicly deposited whole genome sequences in the GISAID database.^26^ For a given test, which is characterized by a specific combination of state and twice-monthly interval, this variable was denoted as (i) Alpha if the prevalence of Alpha variant sequences was > 0.5, (ii) Delta if the prevalence of Delta variant sequences was > 0.5, (ii) neither if the prevalence of both Alpha and Delta variant sequences was < 0.5, or (iv) Unknown if there were fewer than 50 sequences deposited.
2. X_10_: Community level exposure risk, categorized into quintiles. For a given county on a given day of testing, community exposure risk was proxied by the trailing 7-day average COVID-19 incidence (in cases per 100,000 individuals).^27^
3. X_11_: Calendar time of full vaccination for the individual who underwent the symptomatic test, categorized in two-week intervals.

The CLR model was then defined by the equation, 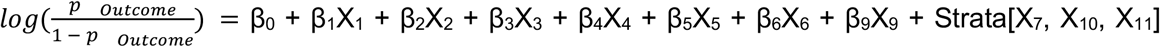, where the covariates and conditioning variables X_1_-X_7_ are the same as described above for the primary analysis. This equation mirrors that used in the primary analysis, except that X_9_ (dominant SARS-CoV-2 variant) was added as a covariate, and the stratifying variable X_8_ (calendar time of test) was replaced with X_10_ (community level exposure risk, categorized into quintiles) and X_11_ (calendar time of vaccination).

It should be noted that in this analysis, the inclusion criteria was modified to include individuals who received their first vaccine dose on or after January 1, 2021. Most individuals vaccinated during January were in high-risk groups for exposure to SARS-CoV-2 or developing severe COVID-19, and so these groups were intentionally excluded from the previous analyses (start date of February 1, 2021). Here, by stratifying on the calendar time of vaccination, we were able to more directly compare these potentially high-risk individuals to each other rather than to likely lower-risk individuals who received their vaccine doses later during the rollout.

## Results

### Primary analysis: change in the odds of symptomatic infection over time after full vaccination

Of 70,583 individuals who received two doses of BNT162b2 18-28 days apart with no evidence of SARS-CoV-2 infection before reaching their date of full vaccination, 11,606 subsequently underwent symptomatic testing and were included in the primary test-negative case-control analysis (**Figure 1, Figure S1**). There were 670 individuals who presented with positive tests (eligible cases) and 12,266 total negative tests (eligible controls) from 10,993 individuals (**Figure S1, Table 1**). Cases and controls were generally similar in age, sex, race, ethnicity, and Elixhauser comorbidity scores to the underlying population of fully vaccinated individuals, but positive tests were disproportionately contributed from Florida (**Table 1**). For the primary analysis, there were 652 cases and 5,946 controls that contributed to analyzable strata (i.e., strata with at least one case and at least one control) (**Figure S1**).

Adjusted for age, sex, race, ethnicity, Elixhauser score, county, and the calendar date of testing, the odds of symptomatic infection were higher at later time points after full vaccination (Odds Ratio [OR]_30 Days_: 1.81, 95% CI: 0.68-4.82; OR_60 Days_: 2.32, 95% CI: 0.97-5.52; OR_90 Days_: 3.5, 95% CI: 1.47-8.35; OR_120 Days_: 3.21, 95% CI: 1.33-7.74) (**Figure 2, Table 2**). Age, race, ethnicity, sex, and Elixhauser score were not significantly associated with the odds of symptomatic infection after full vaccination (**Table 2**). However, the odds of non-COVID-19 hospitalization (the negative control outcome) decreased significantly with time since BNT162b2 vaccination (**Figure 2, Table 2**), suggesting a source of confounding in the design which could lead to underestimation of a waning effect if it is truly present.

**Table 2.**
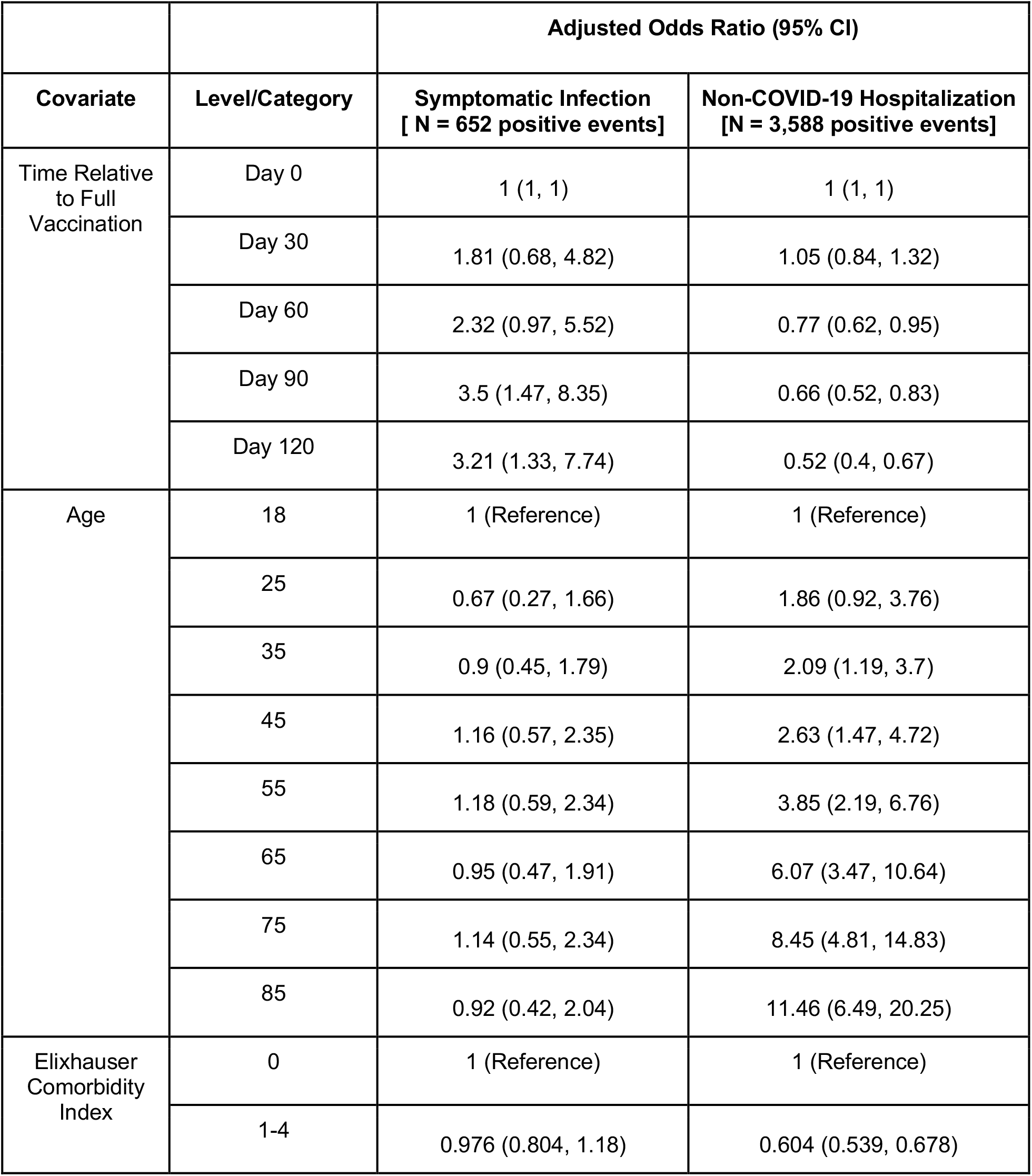
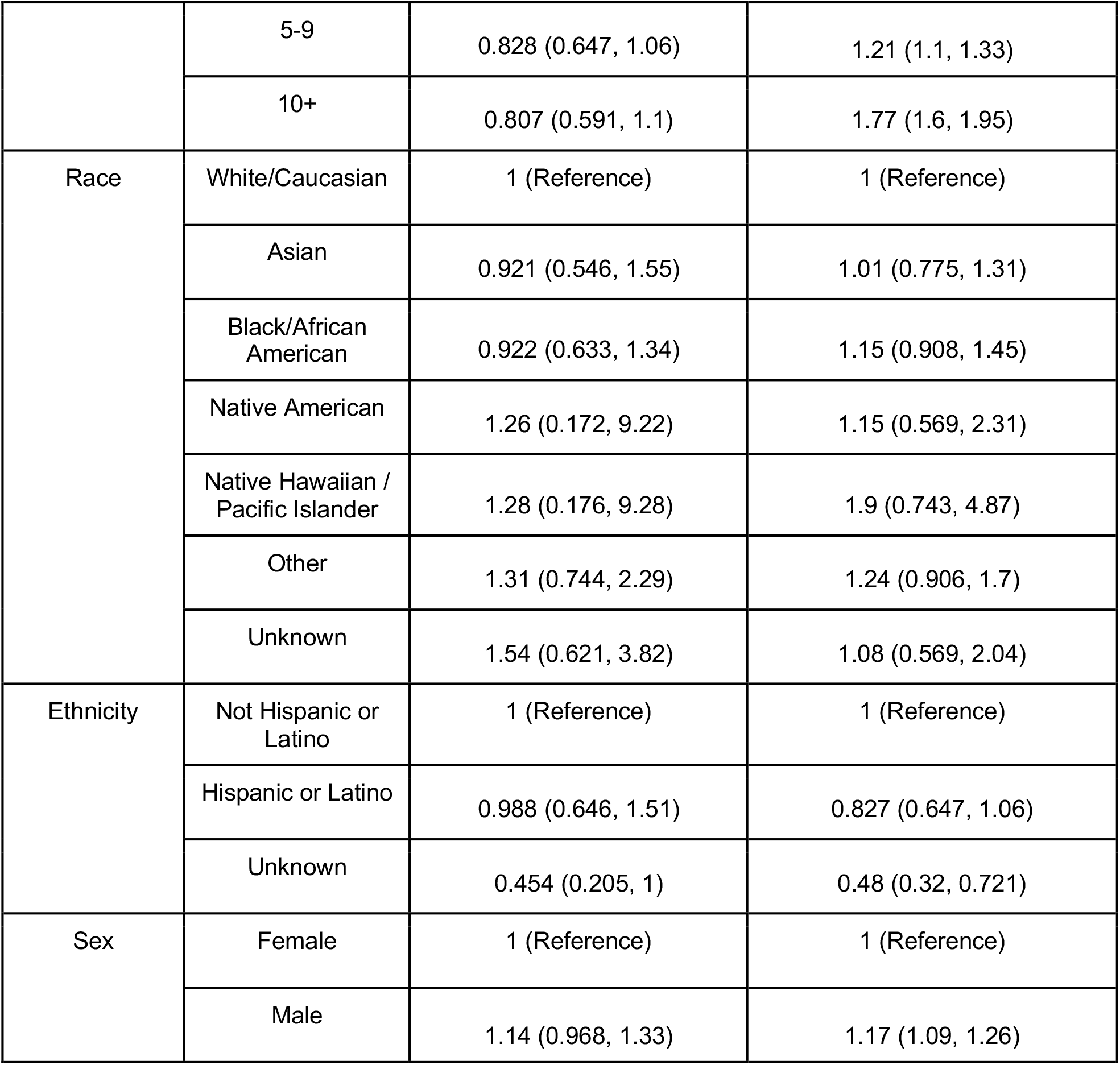
Primary analysis: adjusted odds of symptomatic SARS-CoV-2 infection and non-COVID-19 hospitalization for the main exposure variable (time since full vaccination) and all other covariates. In total, there were 652 positive symptomatic tests and 3,588 non-COVID-19 hospitalizations which contributed to analyzable strata. Adjusted odds were estimated with linear spline equations (for time relative to full vaccination and age) or by exponentiating the coefficients derived from conditional logistic regression models fit separately for each outcome. For “Time Relative to Full Vaccination”, knots through the 90th percentile of the days since full vaccination for the entire cohort are included.

| Covariate | Level/Category | Adjusted Odds Ratio (95% CI) |  |
| --- | --- | --- | --- |
|  |  | Symptomatic Infection<br>[ N = 652 positive events] | Non-COVID-19 Hospitalization<br>[N = 3,588 positive events] |
| Time Relative to Full Vaccination | Day 0 | 1 (1, 1) | 1 (1, 1) |
|  | Day 30 | 1.81 (0.68, 4.82) | 1.05 (0.84, 1.32) |
|  | Day 60 | 2.32 (0.97, 5.52) | 0.77 (0.62, 0.95) |
|  | Day 90 | 3.5 (1.47, 8.35) | 0.66 (0.52, 0.83) |
|  | Day 120 | 3.21 (1.33, 7.74) | 0.52 (0.4, 0.67) |
| Age | 18 | 1 (Reference) | 1 (Reference) |
|  | 25 | 0.67 (0.27, 1.66) | 1.86 (0.92, 3.76) |
|  | 35 | 0.9 (0.45, 1.79) | 2.09 (1.19, 3.7) |
|  | 45 | 1.16 (0.57, 2.35) | 2.63 (1.47, 4.72) |
|  | 55 | 1.18 (0.59, 2.34) | 3.85 (2.19, 6.76) |
|  | 65 | 0.95 (0.47, 1.91) | 6.07 (3.47, 10.64) |
|  | 75 | 1.14 (0.55, 2.34) | 8.45 (4.81, 14.83) |
|  | 85 | 0.92 (0.42, 2.04) | 11.46 (6.49, 20.25) |
| Elixhauser Comorbidity Index | 0 | 1 (Reference) | 1 (Reference) |
|  | 1-4 | 0.976 (0.804, 1.18) | 0.604 (0.539, 0.678) |
|  | 5-9 | 0.828 (0.647, 1.06) | 1.21 (1.1, 1.33) |
|  | 10+ | 0.807 (0.591, 1.1) | 1.77 (1.6, 1.95) |
| Race | White/Caucasian | 1 (Reference) | 1 (Reference) |
|  | Asian | 0.921 (0.546, 1.55) | 1.01 (0.775, 1.31) |
|  | Black/African American | 0.922 (0.633, 1.34) | 1.15 (0.908, 1.45) |
|  | Native American | 1.26 (0.172, 9.22) | 1.15 (0.569, 2.31) |
|  | Native Hawaiian / Pacific Islander | 1.28 (0.176, 9.28) | 1.9 (0.743, 4.87) |
|  | Other | 1.31 (0.744, 2.29) | 1.24 (0.906, 1.7) |
|  | Unknown | 1.54 (0.621, 3.82) | 1.08 (0.569, 2.04) |
| Ethnicity | Not Hispanic or Latino | 1 (Reference) | 1 (Reference) |
|  | Hispanic or Latino | 0.988 (0.646, 1.51) | 0.827 (0.647, 1.06) |
|  | Unknown | 0.454 (0.205, 1) | 0.48 (0.32, 0.721) |
| Sex | Female | 1 (Reference) | 1 (Reference) |
|  | Male | 1.14 (0.968, 1.33) | 1.17 (1.09, 1.26) |

**Figure 2.**
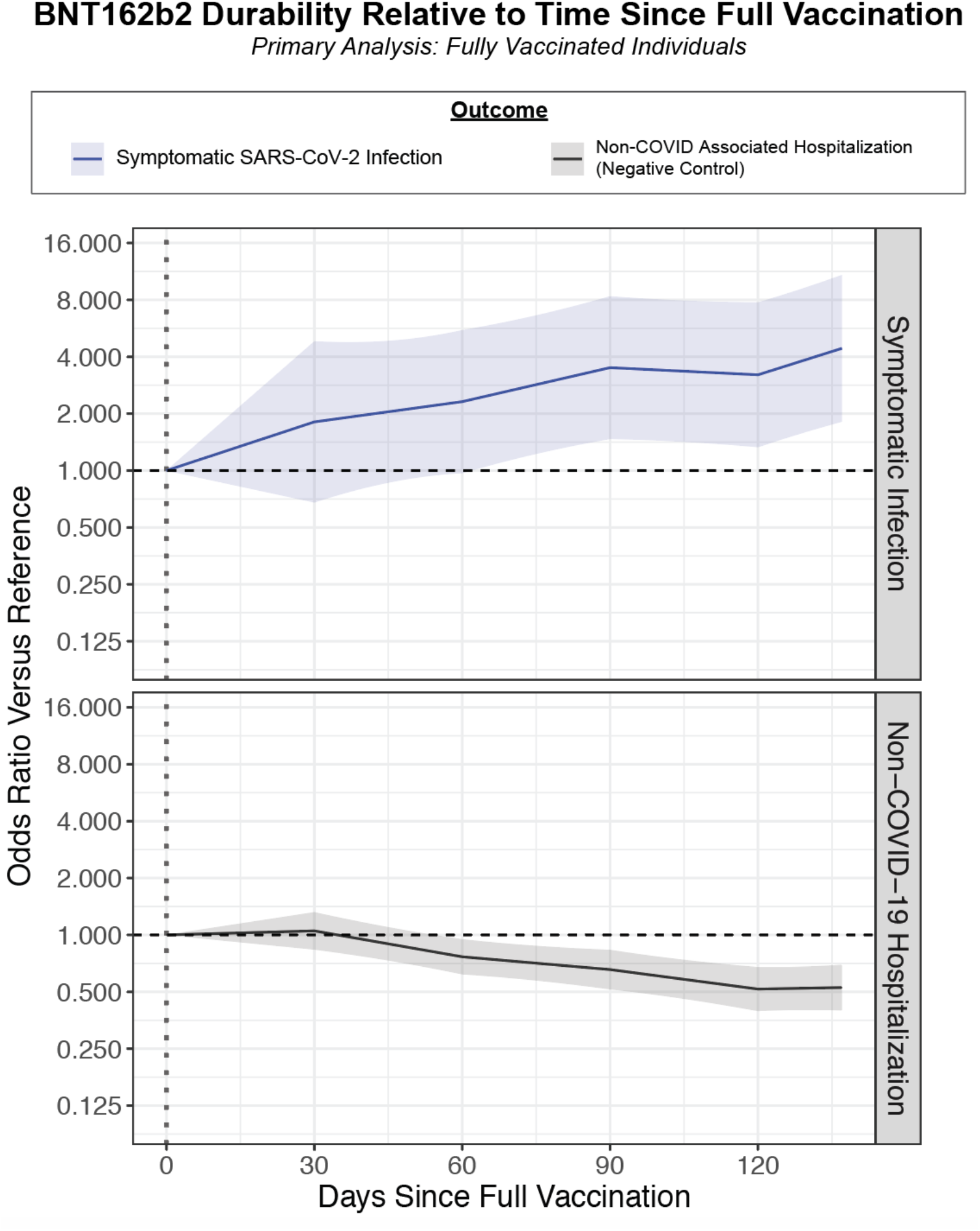
Primary analysis: relationship between time since full vaccination and the adjusted odds of experiencing each outcome of interest. The outcomes are symptomatic SARS-CoV-2 infection (blue; 652 cases and 6,256 controls) and non-COVID-19 hospitalization (gray; 3,588 cases and 7,623 controls). Each curve indicates the adjusted odds ratio comparing the odds of experiencing the outcome at the given time compared to at the time of full vaccination (Day 0 on this plot), which is expected to correspond to maximal vaccine-mediated protection. The adjusted odds ratios at each day are calculated from the linear spline equations, and data is shown through the 90th percentile of the “Days Since Full Vaccination” for the entire cohort. The shaded region indicates the 95% confidence interval of the odds ratio. Related to **Table 2**.

### Secondary analysis: change in the odds of infection over time after the first vaccine dose

Of 86,349 individuals who received at least one dose of BNT162b2 with no positive SARS-CoV-2 tests prior to vaccination, 15,180 underwent symptomatic testing after their first dose. There were 1,061 individuals who presented as positive cases (391 before and 670 after expected full vaccination, respectively) and 16,165 negative tests (3,798 before and 12,367 after expected full vaccination, respectively) from 14,205 individuals (**Table 3**). Compared to four days after the first dose (a proxy for the unvaccinated state), the odds of symptomatic infection decreased through the expected second dose and full vaccination dates (e.g., OR_Day 21_: 0.48; 95% CI: 0.30-0.77; OR_Day 35_: 0.20; 95% CI: 0.12-0.34), corresponding to the onset of vaccine effectiveness (**Figure 3, Table 4**). The odds at time points further removed from the first dose were higher than than those at the expected full vaccination date but importantly remained lower than those at the proxy unvaccinated state (e.g., OR_Day 120_: 0.31; 95% CI: 0.20-0.48; OR_Day 150_: 0.3; 95% CI: 0.19-0.45) (**Figure 3, Table 4**).

**Table 3.**
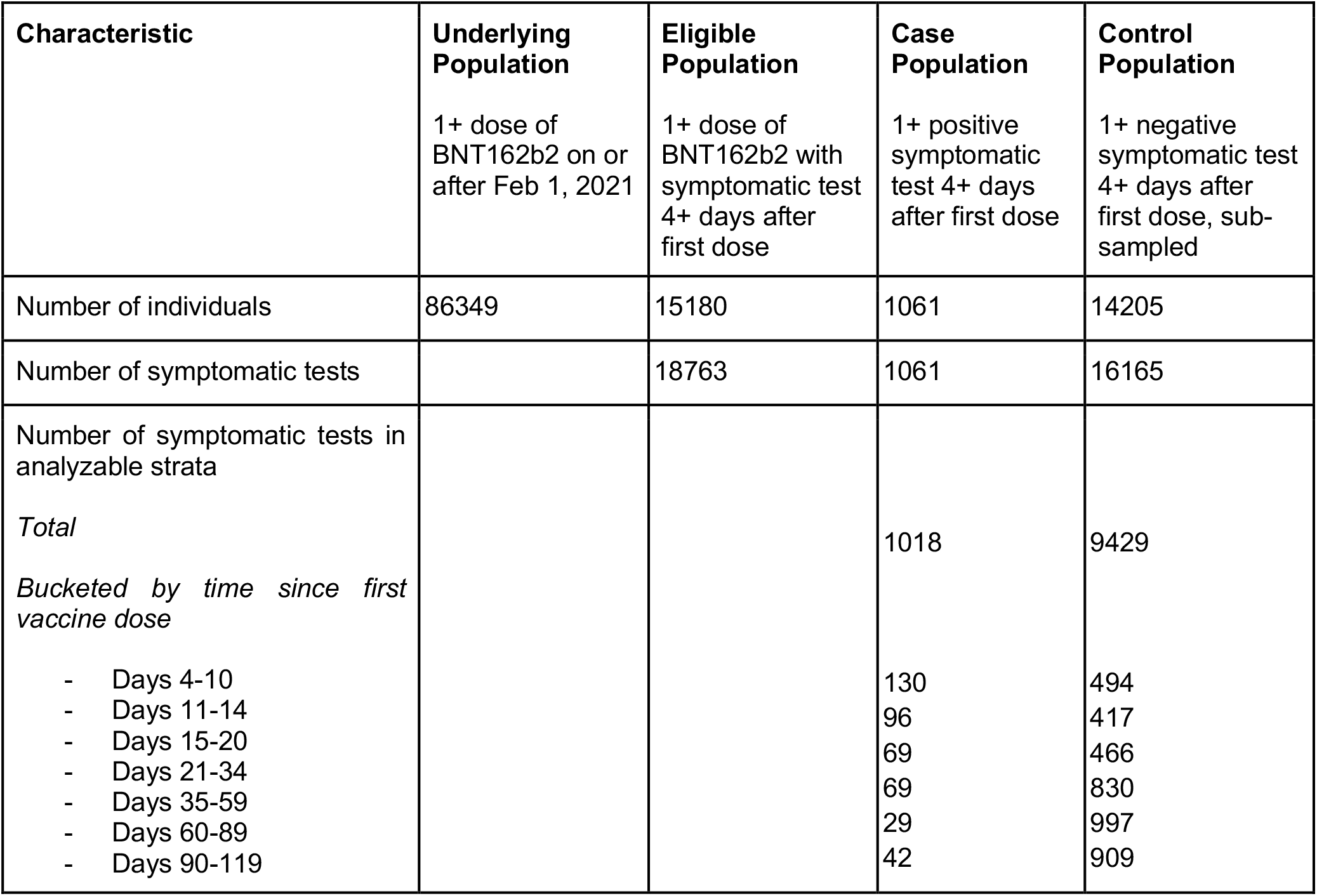
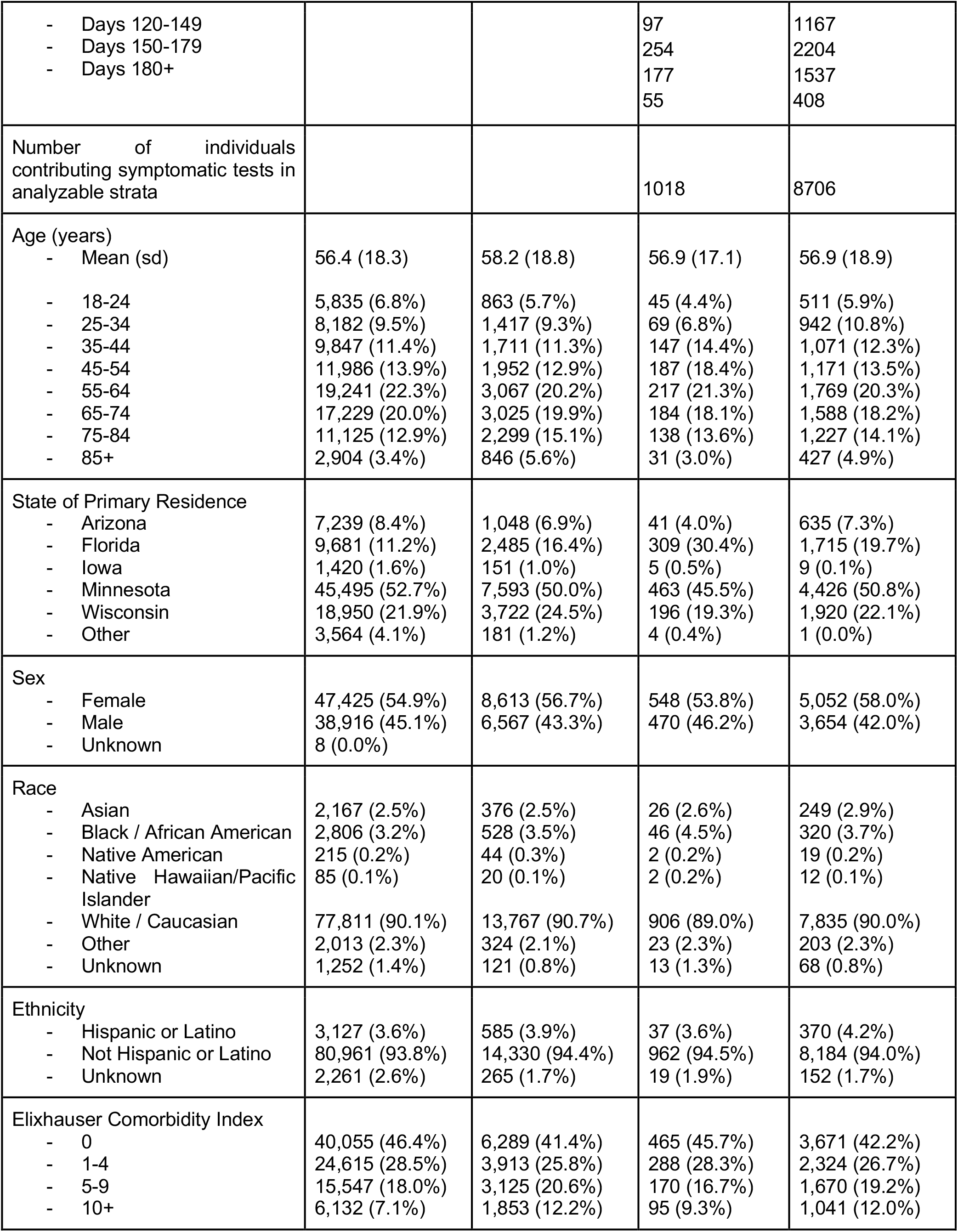
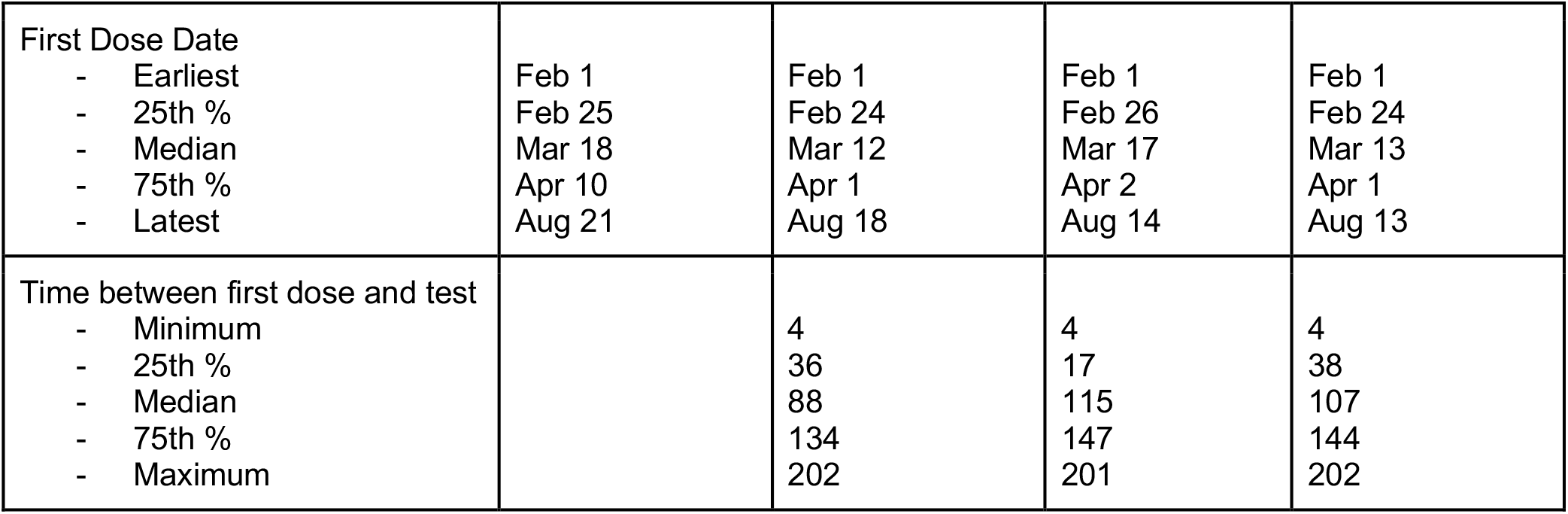
Demographic and clinical characteristics of cases and controls for the secondary analysis of BNT162b2 starting after the first vaccine dose. The underlying population corresponds to the set of individuals who received their first BNT162b2 dose on or after February 1, 2021, had not received any prior doses of COVID-19 vaccines, and had no record of a positive PCR test prior to the first dose of BNT162b2. The eligible population corresponds to the subset of the underlying population who underwent at least one symptomatic test four or more days after their first dose. Cases correspond to the first positive symptomatic test for a given individual in the eligible population; by definition, the number of individuals contributing cases is the same as the number of cases. Controls correspond to negative symptomatic tests after which occur before the given individual has experienced any positive SARS-CoV-2 PCR tests; an individual can contribute multiple controls during the study period, and so the number of individuals in the control population is less than the total number of tests (controls) contributed. Because an individual can contribute negative tests (controls) prior to contributing a positive test, the number of individuals in the eligible population is smaller than the sum of the number of individuals in the case and control populations. Sub-sampling in the control population refers to the process in which negative tests from a given individual were (i) excluded if they occurred after a positive test or within the 15 days prior to a positive test (possible false negative), (ii) randomly sampled if they occurred within 15 days of each other (possibly during the same symptomatic illness), and (iii) randomly sampled if the individual contributed more than three negative tests during the study period. A stratum (defined by the regression equation as a unique combination of county and calendar week of testing) is considered analyzable if it includes at least one case and at least one control, because strata including only cases or only controls do not contribute to the estimation of the regression coefficients. For cases and controls, all summarized characteristics correspond to only individuals who contributed at least one symptomatic test to an analyzable stratum.

| Characteristic | Underlying Population | Eligible Population | Case Population | Control Population |
| --- | --- | --- | --- | --- |
|  | 1+ dose of BNT162b2 on or after Feb 1, 2021 | 1+ dose of BNT162b2 with symptomatic test 4+ days after first dose | 1+ positive symptomatic test 4+ days after first dose | 1+ negative symptomatic test 4+ days after first dose, sub-sampled |
| Number of individuals | 86349 | 15180 | 1061 | 14205 |
| Number of symptomatic tests |  | 18763 | 1061 | 16165 |
| Number of symptomatic tests in analyzable strata |  |  |  |  |
| *Total* |  |  | 1018 | 9429 |
| *Bucketed by time since first vaccine dose* |  |  |  |  |
| - Days 4-10 |  |  | 130 | 494 |
| - Days 11-14 |  |  | 96 | 417 |
| - Days 15-20 |  |  | 69 | 466 |
| - Days 21-34 |  |  | 69 | 830 |
| - Days 35-59 |  |  | 29 | 997 |
| - Days 60-89 |  |  | 42 | 909 |
| - Days 90-119 |  |  |  |  |
| - Days 120-149<br>- Days 150-179<br>- Days 180+ |  |  | 97<br>254<br>177<br>55 | 1167<br>2204<br>1537<br>408 |
| Number of individuals contributing symptomatic tests in analyzable strata |  |  | 1018 | 8706 |
| Age (years) |  |  |  |  |
| - Mean (sd) | 56.4 (18.3) | 58.2 (18.8) | 56.9 (17.1) | 56.9 (18.9) |
| - 18-24<br>- 25-34<br>- 35-44<br>- 45-54<br>- 55-64<br>- 65-74<br>- 75-84<br>- 85+ | 5,835 (6.8%)<br>8,182 (9.5%)<br>9,847 (11.4%)<br>11,986 (13.9%)<br>19,241 (22.3%)<br>17,229 (20.0%)<br>11,125 (12.9%)<br>2,904 (3.4%) | 863 (5.7%)<br>1,417 (9.3%)<br>1,711 (11.3%)<br>1,952 (12.9%)<br>3,067 (20.2%)<br>3,025 (19.9%)<br>2,299 (15.1%)<br>846 (5.6%) | 45 (4.4%)<br>69 (6.8%)<br>147 (14.4%)<br>187 (18.4%)<br>217 (21.3%)<br>184 (18.1%)<br>138 (13.6%)<br>31 (3.0%) | 511 (5.9%)<br>942 (10.8%)<br>1,071 (12.3%)<br>1,171 (13.5%)<br>1,769 (20.3%)<br>1,588 (18.2%)<br>1,227 (14.1%)<br>427 (4.9%) |
| State of Primary Residence |  |  |  |  |
| - Arizona<br>- Florida<br>- Iowa<br>- Minnesota<br>- Wisconsin<br>- Other | 7,239 (8.4%)<br>9,681 (11.2%)<br>1,420 (1.6%)<br>45,495 (52.7%)<br>18,950 (21.9%)<br>3,564 (4.1%) | 1,048 (6.9%)<br>2,485 (16.4%)<br>151 (1.0%)<br>7,593 (50.0%)<br>3,722 (24.5%)<br>181 (1.2%) | 41 (4.0%)<br>309 (30.4%)<br>5 (0.5%)<br>463 (45.5%)<br>196 (19.3%)<br>4 (0.4%) | 635 (7.3%)<br>1,715 (19.7%)<br>9 (0.1%)<br>4,426 (50.8%)<br>1,920 (22.1%)<br>1 (0.0%) |
| Sex |  |  |  |  |
| - Female<br>- Male<br>- Unknown | 47,425 (54.9%)<br>38,916 (45.1%)<br>8 (0.0%) | 8,613 (56.7%)<br>6,567 (43.3%) | 548 (53.8%)<br>470 (46.2%) | 5,052 (58.0%)<br>3,654 (42.0%) |
| Race |  |  |  |  |
| - Asian<br>- Black / African American<br>- Native American<br>- Native Hawaiian/Pacific Islander<br>- White / Caucasian<br>- Other<br>- Unknown | 2,167 (2.5%)<br>2,806 (3.2%)<br>215 (0.2%)<br>85 (0.1%)<br>77,811 (90.1%)<br>2,013 (2.3%)<br>1,252 (1.4%) | 376 (2.5%)<br>528 (3.5%)<br>44 (0.3%)<br>20 (0.1%)<br>13,767 (90.7%)<br>324 (2.1%)<br>121 (0.8%) | 26 (2.6%)<br>46 (4.5%)<br>2 (0.2%)<br>2 (0.2%)<br>906 (89.0%)<br>23 (2.3%)<br>13 (1.3%) | 249 (2.9%)<br>320 (3.7%)<br>19 (0.2%)<br>12 (0.1%)<br>7,835 (90.0%)<br>203 (2.3%)<br>68 (0.8%) |
| Ethnicity |  |  |  |  |
| - Hispanic or Latino<br>- Not Hispanic or Latino<br>- Unknown | 3,127 (3.6%)<br>80,961 (93.8%)<br>2,261 (2.6%) | 585 (3.9%)<br>14,330 (94.4%)<br>265 (1.7%) | 37 (3.6%)<br>962 (94.5%)<br>19 (1.9%) | 370 (4.2%)<br>8,184 (94.0%)<br>152 (1.7%) |
| Elixhauser Comorbidity Index |  |  |  |  |
| - 0<br>- 1-4<br>- 5-9<br>- 10+ | 40,055 (46.4%)<br>24,615 (28.5%)<br>15,547 (18.0%)<br>6,132 (7.1%) | 6,289 (41.4%)<br>3,913 (25.8%)<br>3,125 (20.6%)<br>1,853 (12.2%) | 465 (45.7%)<br>288 (28.3%)<br>170 (16.7%)<br>95 (9.3%) | 3,671 (42.2%)<br>2,324 (26.7%)<br>1,670 (19.2%)<br>1,041 (12.0%) |
| First Dose Date<br>- Earliest<br>- 25th %<br>- Median<br>- 75th %<br>- Latest | Feb 1<br>Feb 25<br>Mar 18<br>Apr 10<br>Aug 21 | Feb 1<br>Feb 24<br>Mar 12<br>Apr 1<br>Aug 18 | Feb 1<br>Feb 26<br>Mar 17<br>Apr 2<br>Aug 14 | Feb 1<br>Feb 24<br>Mar 13<br>Apr 1<br>Aug 13 |
| Time between first dose and test<br>- Minimum<br>- 25th %<br>- Median<br>- 75th %<br>- Maximum |  | 4<br>36<br>88<br>134<br>202 | 4<br>17<br>115<br>147<br>201 | 4<br>38<br>107<br>144<br>202 |

**Table 4.** Secondary analysis: adjusted odds of symptomatic SARS-CoV-2 infection and non-COVID-19 hospitalization by time since first BNT162b2 dose. There were 364 positive symptomatic tests before the expected date of full vaccination and 654 positive symptomatic tests after the expected date of full vaccination which contributed to analyzable strata. There were 872 non-COVID-19 hospitalizations before the expected date of full vaccination and 3,677 non-COVID-19 hospitalizations after the date of full vaccination which contributed to analyzable strata. Adjusted odds were estimated with linear spline equations derived from conditional logistic regression models fit separately for each outcome. For “Time Relative to First Vaccine Dose”, knots through the 90th percentile of the days since first vaccination for the entire cohort are included.

| Time Relative to First Vaccine Dose | Adjusted Odds Ratio (95% CI) |  |
| --- | --- | --- |
|  | Symptomatic Infection<br>[N = 1,018 positive events] | Non-COVID-19 Hospitalization<br>[N = 4,549 positive events] |
| Day 4 | 1<br>(Reference) | 1<br>(Reference) |
| Days 10 | 0.86 (0.53, 1.4) | 0.83 (0.55, 1.25) |
| Day 15 | 0.71 (0.46, 1.09) | 0.95 (0.67, 1.33) |
| Day 21<br>(Expected second dose) | 0.48 (0.3, 0.77) | 0.81 (0.58, 1.12) |
| Day 35<br>(Expected full vaccination) | 0.2 (0.12, 0.34) | 0.94 (0.68, 1.29) |
| Day 60 | 0.16 (0.09, 0.28) | 0.94 (0.69, 1.29) |
| Day 90 | 0.24 (0.14, 0.4) | 0.74 (0.53, 1.02) |
| Day 120 | 0.31 (0.2, 0.48) | 0.6 (0.43, 0.84) |
| Day 150 | 0.3 (0.19, 0.45) | 0.5 (0.35, 0.7) |

**Figure 3.**
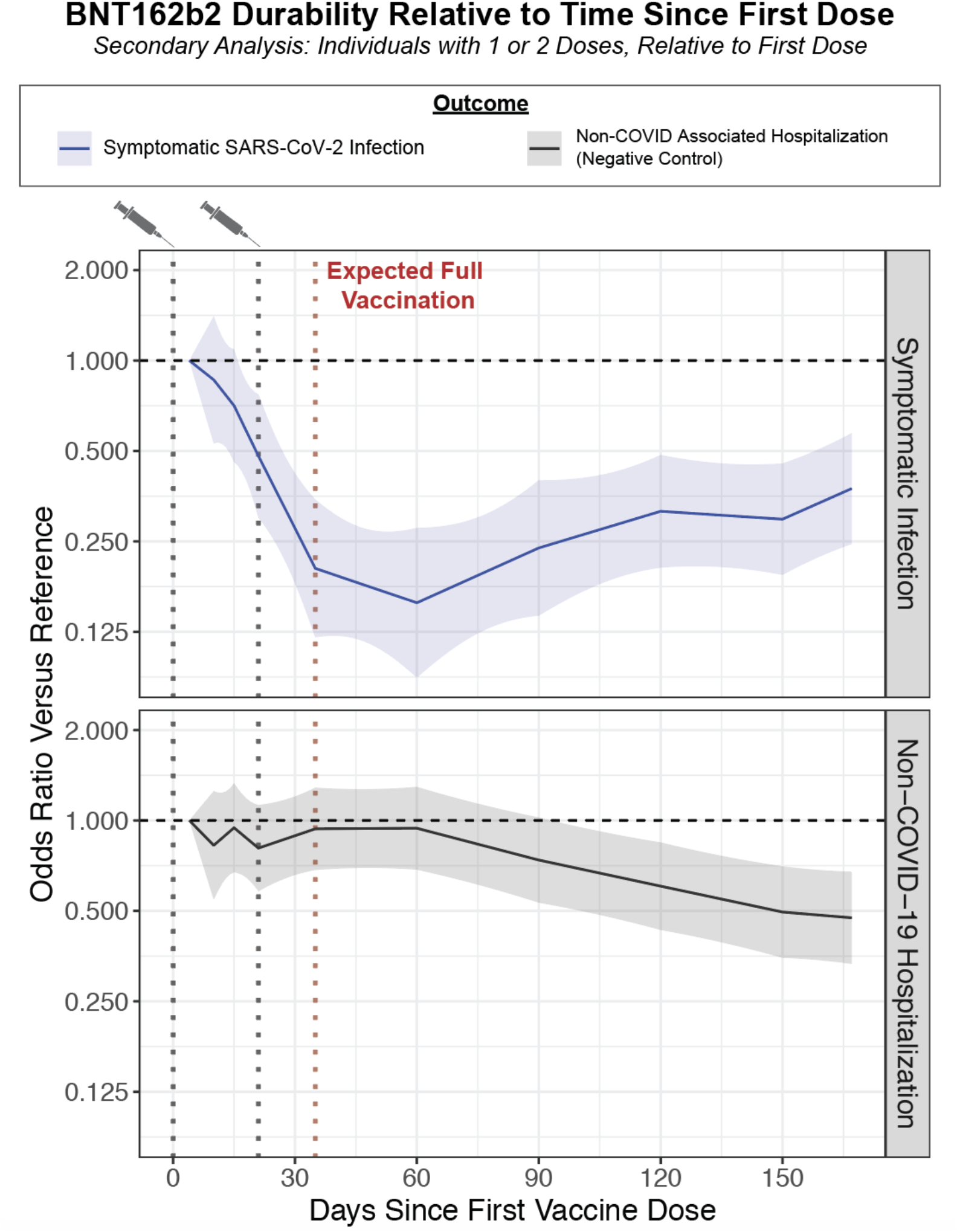
Secondary analysis: relationship between time since first vaccine dose and the adjusted odds of experiencing each outcome of interest. The outcomes are symptomatic SARS-CoV-2 infection (blue; 364 and 654 cases before and after full vaccination) and non-COVID-19 hospitalization (gray; 872 and 3,677 cases before and after full vaccination). Each curve indicates the adjusted odds ratio comparing the odds of experiencing the outcome at the given time versus at four days after the first dose (“reference”), which is expected to approximate an unvaccinated state. The adjusted odds ratios at each day are calculated from the linear spline equations, and data is shown through the 90th percentile of the “Days Since First Vaccine Dose” for the entire cohort. The shaded region indicates the 95% confidence interval of the odds ratio. Related to **Table 4**.

### Sensitivity analysis 1: Age subgroup analysis

Among individuals aged 18-44, 45-64, and 65+ years old, there were 247, 385, and 344 positive symptomatic tests (cases) versus 1950, 2173, and 2376 negative symptomatic tests (controls) after the first BNT162b2 dose. Subgroup analyses suggest that there are indeed trends of waning protection against symptomatic infection after full vaccination for all three groups (**Figure S2, Table S1**). Importantly, in each group, the odds of symptomatic infection were still significantly lower 150 days after the first dose compared to four days after the first dose, with odds ratios of 0.37 (95% CI: 0.17-0.81), 0.27 (95% CI: 0.13-0.54), and 0.18 (95% CI: 0.06-0.5) for the 18-44, 45-64, and 65+ groups, respectively (**Figure S2, Table S1**).

### Sensitivity analysis 2: Stratification on time of vaccination rather than time of testing

One source of confounding in our primary analysis, which could contribute to the unexpected negative control findings described previously, is that any given stratum may include individuals who became eligible or chose to get vaccinated at different times. To address this, we modified the CLR to instead stratify on the time of vaccination and the county-level COVID-19 incidence at the time of PCR testing (see **Methods**). Among 97,487 individuals who received their first dose on or after January 1, 2021 and were at risk for infection at their date of full vaccination, there were subsequently 974 positive symptomatic tests (cases) and 11,371 negative symptomatic tests (controls) contributing to analyzable strata (**Table S2**). With this modified approach, the odds of non-COVID-19 hospitalization (negative control outcome) were no longer associated with time since BNT162b2 vaccination (**Figure S3, Table S3**). There was a stronger signal for waning immunity than in the primary analysis, as the odds of symptomatic infection during the 120 and 150 days after full vaccination were 7.25 (95% CI: 3.47-15.2) and 10.3 (95% CI: 5.03-21.2) times higher than at the date of full vaccination, respectively (**Figure S3, Table S3**).

## Discussion

Taken together, these data show that the risk of symptomatic infection several months after BNT162b2 vaccination is higher than at the date of full vaccination, but the risk of infection remains significantly lower than at a baseline or unvaccinated state, supporting public health recommendations for universal vaccination. These data constitute an early signal for waning protection against symptomatic infection conferred by vaccination with BNT162b2. Critically, we were not adequately powered to assess the durability of protection against severe COVID-19 (e.g. hospitalization, ICU admission, and death) as these events were fortunately rare in all time periods for vaccinated individuals. We acknowledge that these outcomes are of primary importance, and it is critical that follow-up studies are performed to answer this question as more cases accumulate.

Importantly, these data do not indicate a complete loss of effectiveness against symptomatic infection over the duration of the study. Instead, the adjusted odds of experiencing a symptomatic infection remain lower 150 days after full vaccination compared to 4 days after the first dose, when immunity more closely approximates the unvaccinated state. This suggests that significant protection against symptomatic infection does persist for months after vaccination. Anecdotally, this is consistent with what has been observed clinically at our sites. Since the beginning of June, over 80% of patients admitted to the hospital with symptomatic COVID-19 are unvaccinated despite less than 40% of Minnesota residents remaining unvaccinated over this period (data not presented).^28^ Further, fewer than 0.5% of over 200,000 individuals recorded as fully vaccinated at Mayo Clinic during the study period have experienced a PCR-confirmed symptomatic breakthrough infection, emphasizing how low the rate of symptomatic infection in vaccinated persons remains overall. Vaccination appears to provide a persistent reduction in risk over time, despite the early evidence of waning protection presented here.

These findings motivate further investigation of strategies to prolong vaccine induced immunity, such as combining vaccination with nonpharmaceutical interventions or administering additional doses of vaccines to vulnerable populations.^20,29^ We emphasize the need to collect additional prospective clinical data to inform recommendations about booster doses, particularly regarding their safety, immunogenicity, and effectiveness against severe outcomes.

It is noteworthy that several reports have suggested waning effectiveness of BNT162b2 against symptomatic infection over time.^12–19^ In line with this, recent studies have demonstrated that antibody titers decline over time after full vaccination with BNT162b2, which is particularly relevant because neutralizing antibody levels are suggested to be highly predictive of protection against SARS-CoV-2 infection.^30^. In one study, the levels of Spike protein antibodies declined by approximately two-fold between 21-41 days and 70+ days after the second dose.^31^ In a separate study of healthcare workers, the levels of neutralizing antibodies and antibodies that specifically recognize the Spike protein receptor binding domain significantly declined over several months after full vaccination.^32^

## Study Limitations

There are several limitations of this study. First, the demographic composition of the studied cohort is not representative of the United States or global population (e.g. over 90% Caucasian). Future investigation should test whether these results apply to more diverse and representative populations. Second, there are individual-level SARS-CoV-2 exposure risk factors and non-pharmaceutical interventions which could not be accounted for in our regression analyses such as occupational risk and adherence to masking and social distancing guidelines. Third, the use of a test-negative study design makes it difficult to assess the durability of protection against asymptomatic infection. While the vaccines are primarily intended to reduce symptomatic infection and severe disease, asymptomatic infections comprise a meaningful fraction of cases and can contribute to community transmission.^33–35^ The test-negative design can also be adversely impacted by the variable and sometimes low sensitivity of SARS-CoV-2 PCR tests (estimates ranging from approximately 30-80%), which will likely result in the misclassification of some cases as controls.^36,37^

Next, we note that our negative control (non-COVID-19 hospitalization) decreased with time since BNT162b2 vaccination in the primary analysis. This suggests some confounding, which may owe to differences in factors such as the baseline health consciousness of individuals who were vaccinated in early versus late phases of the rollout or the dynamic nature of elective procedures and non-COVID-19 related healthcare during the pandemic. However, the direction of this confounding would support the overall conclusion (i.e., waning protection against symptomatic infection) while possibly underestimating its magnitude. We attempted to address this through a sensitivity analysis stratifying on the time of vaccination rather than the time of testing. Although this approach is not standard for a case-control study design, the results were indeed consistent with the conclusion of the primary analysis and showed the expected lack of change in the negative control outcome with respect to time since vaccination.

Additionally, this analysis only considered BNT162b2, but other COVID-19 vaccines (e.g., mRNA-1273 and Ad26COV2.S) merit independent evaluation. Because BNT162b2 was available earlier and distributed more widely than other vaccines in the United States, a larger population was available for study at this juncture. As additional breakthrough cases accumulate for the other vaccines, similar studies evaluating their durability will be critical. Finally, due to the rarity of hospitalization, ICU admission, and death in our vaccinated cohorts, we were not able to robustly assess whether protection against these severe outcomes changes over time.

## Conclusions

BNT162b2 demonstrated strong protection against symptomatic and severe disease in clinical trials and the real world setting during early phases of the vaccine rollout internationally.^3,5,8,9,38–41^ We have now entered an era in which the vaccine durability, both over time and in the face of a rapidly evolving landscape of SARS-CoV-2 variants, must be continuously evaluated. This study constitutes an early signal that protection against symptomatic infection wanes over time after BNT162b2 vaccination. It remains critical to continue delivering first and second vaccine doses to as many people as possible, while also considering strategies to boost immunity among at-risk populations.

## Data Availability

After publication, the data will be made available upon reasonable requests to the corresponding authors. A proposal with a detailed description of study objectives and the statistical analysis plan will be needed for evaluation of the reasonability of requests. Deidentified data will be provided after approval from the corresponding authors and the Mayo Clinic.

## Acknowledgements

The authors thank Murali Aravamudan for his feedback on this manuscript.

## Supporting Information

**Figure S1.**
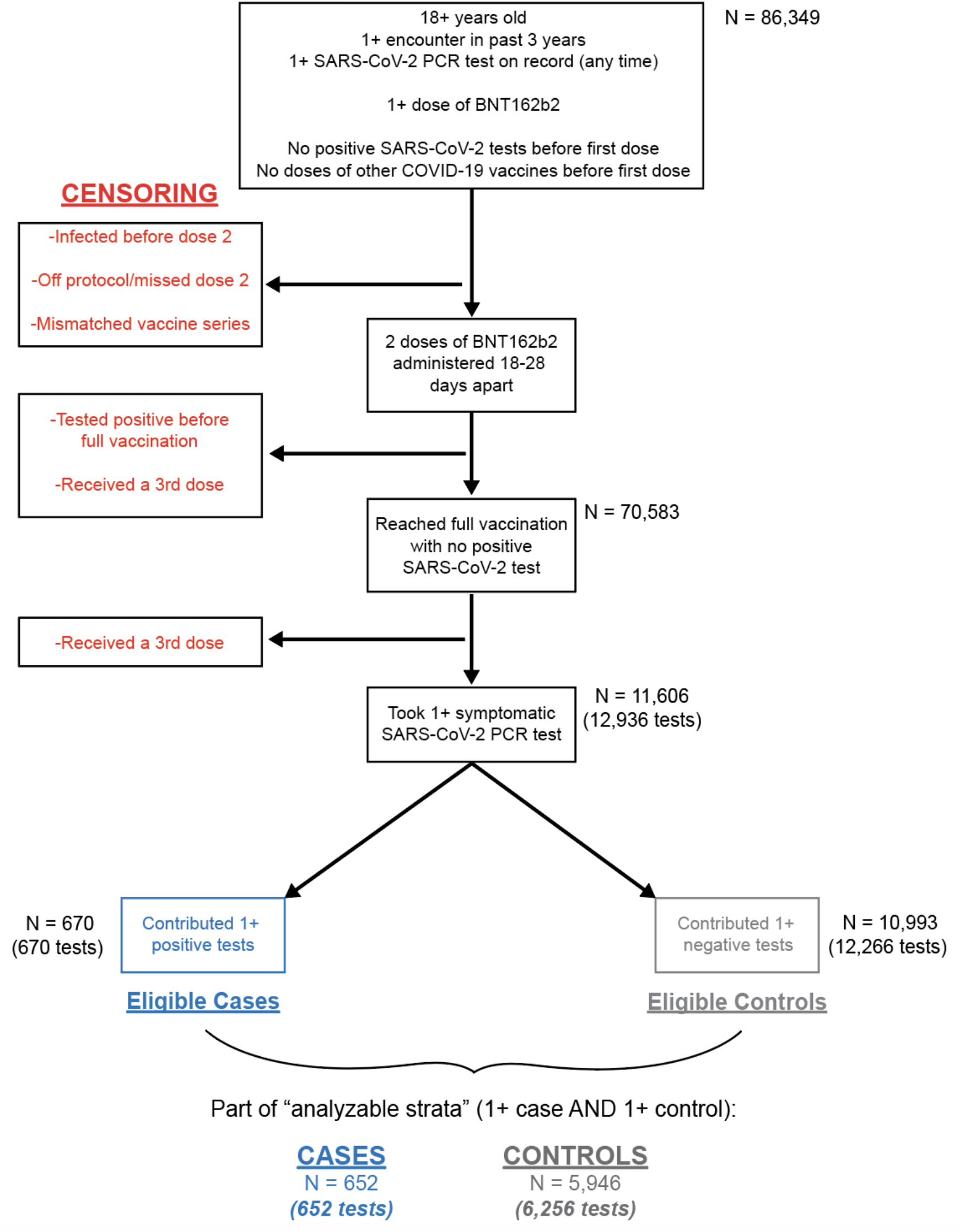
Cohort development flowchart showing derivation of fully vaccinated cohort considered for the primary analysis of BNT162b2 durability.

**Figure S2.**
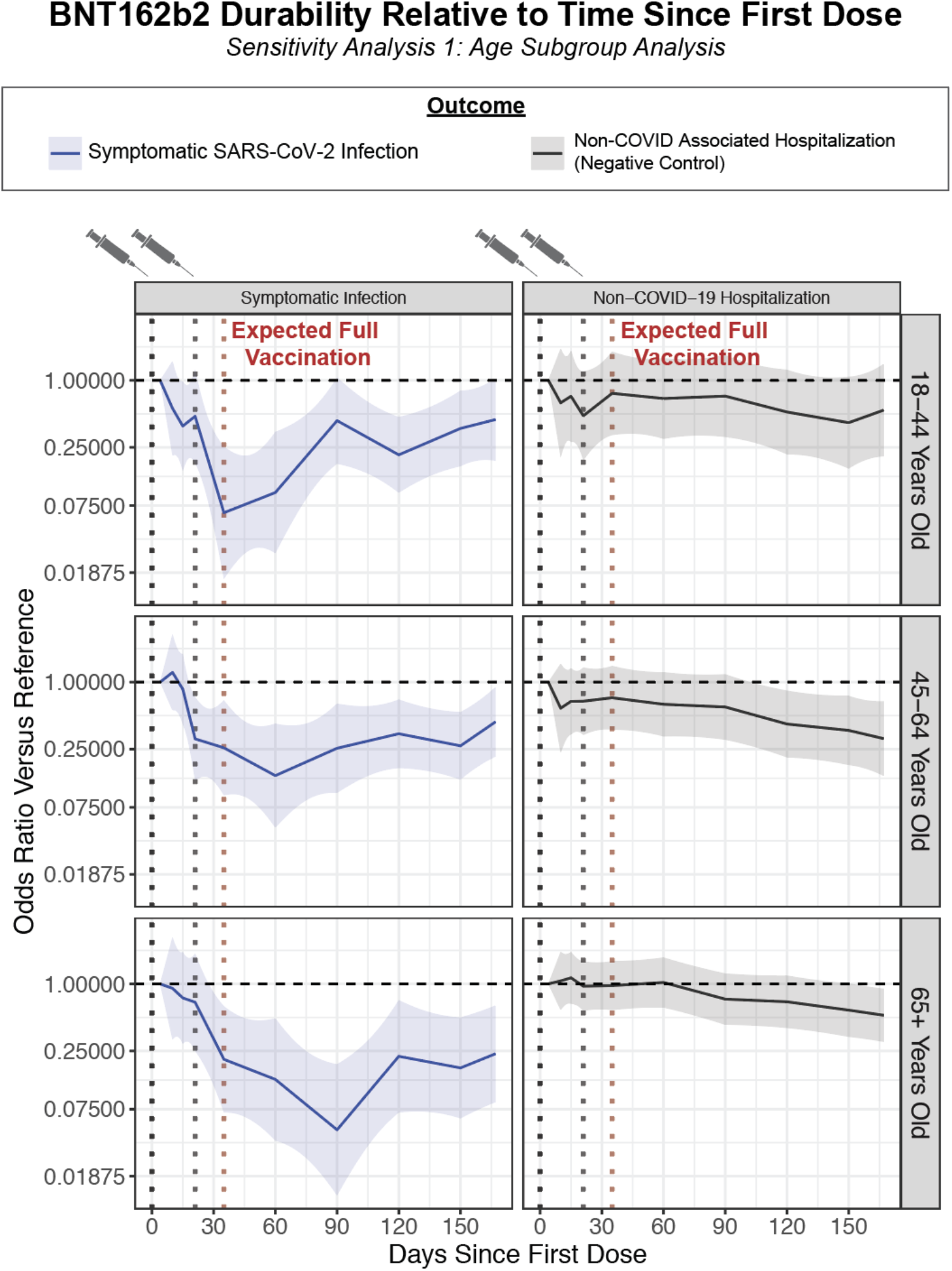
Sensitivity analysis 1: relationship between time since first vaccine dose and the adjusted odds of outcomes of interest in three age subgroups. The outcomes are symptomatic SARS-CoV-2 infection (blue; n = 247 for 18-44; n = 385 for 45-64; n = 344 for 65+) and non-COVID-19 hospitalization (gray; n = 396 for 18-44; n = 1,030 for 45-64; n = 2,821 for 65+). Each curve indicates the adjusted odds ratio comparing the odds of experiencing the outcome at the given time compared to four days after the first dose, which is expected to approximate the unvaccinated state. The adjusted odds ratios at each day are calculated from the linear spline equations, and data is shown through the 90th percentile of the “Days Since First Dose” for the entire cohort. Data is shown for individuals 18-44 (top), 45-64 (middle), and at least 65 (bottom) years old. The shaded region indicates the 95% confidence interval of the odds ratio. Related to **Table S1**.

**Figure S3.**
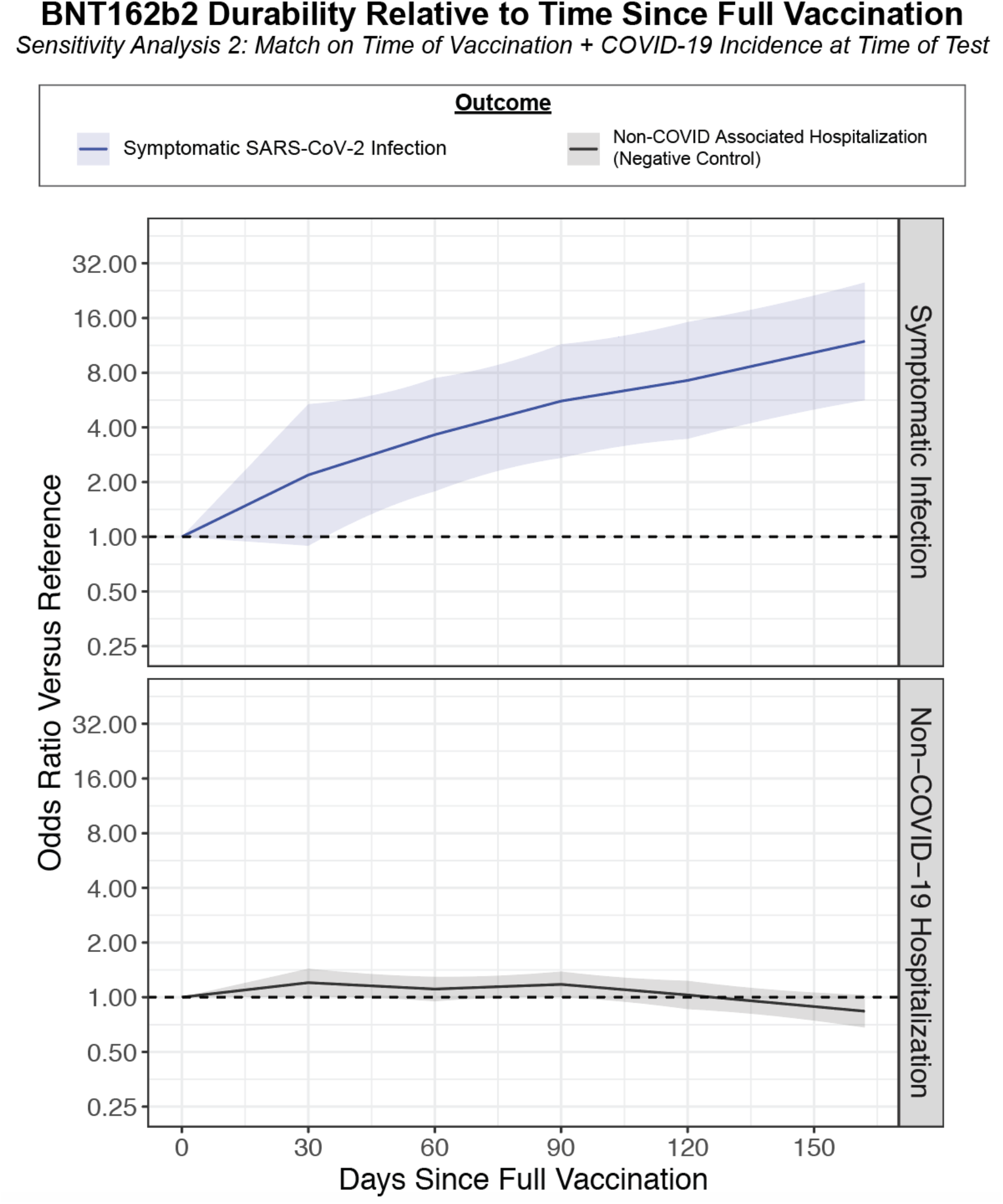
Sensitivity analysis 2: relationship between time since full vaccination and adjusted odds of outcomes of interest with matching on time of vaccination and county-level COVID-19 incidence. The outcomes are symptomatic SARS-CoV-2 infection (blue; 973 cases) and non-COVID-19 hospitalization (gray; 5,249 cases). Each curve indicates the adjusted odds ratio comparing the odds of experiencing the outcome at the given time compared to at the time of full vaccination, which is expected to correspond to maximal vaccine-mediated protection. The adjusted odds ratios at each day are calculated from the linear spline equations, and data is shown through the 90th percentile of the “Days Since Full Vaccination” for the entire cohort. The shaded region indicates the 95% confidence interval of the odds ratio. Related to **Table S2**.

**Table S1.** Sensitivity analysis 1: adjusted odds of symptomatic SARS-CoV-2 infection and non-COVID-19 hospitalization by time since first vaccine dose, split by age group. Adjusted odds were estimated with linear spline equations derived from conditional logistic regression models fit separately for each outcome. The number of positive positive events in each age group is provided in the first column. Related to **Figure S2**. Knots through the 90th percentile of the days since first vaccination for the entire cohort are included.

| Age Group | Time Relative to First Dose | Adjusted Odds Ratio (95% CI) |  |
| --- | --- | --- | --- |
|  |  | Symptomatic Infection | Non-COVID-19 Hospitalization |
| 18-44 years<br>(247 infections;<br>396 hospitalizations) | Day 4 | 1 (Reference) | 1 (Reference) |
|  | Days 10 | 0.56 (0.21, 1.48) | 0.63 (0.21, 1.92) |
|  | Day 15 | 0.42 (0.18, 0.98) | 0.7 (0.29, 1.69) |
|  | Day 21<br>(Expected second dose) | 0.48 (0.18, 1.24) | 0.48 (0.19, 1.2) |
|  | Day 35<br>(Expected full vaccination) | 0.06 (0.02, 0.25) | 0.76 (0.32, 1.81) |
|  | Day 60 | 0.1 (0.03, 0.34) | 0.69 (0.3, 1.58) |
|  | Day 90 | 0.43 (0.18, 1.06) | 0.72 (0.31, 1.68) |
|  | Day 120 | 0.21 (0.1, 0.47) | 0.52 (0.22, 1.24) |
|  | Day 150 | 0.37 (0.17, 0.81) | 0.42 (0.16, 1.07) |
| 45-64 years<br>(385 infections;<br>1,030 hospitalizations) | Day 4 | 1 (Reference) | 1 (Reference) |
|  | Days 10 | 1.22 (0.56, 2.68) | 0.58 (0.23, 1.46) |
|  | Day 15 | 0.93 (0.48, 1.8) | 0.65 (0.33, 1.29) |
|  | Day 21<br>(Expected second dose) | 0.31 (0.13, 0.73) | 0.67 (0.34, 1.35) |
|  | Day 35<br>(Expected full vaccination) | 0.26 (0.1, 0.69) | 0.72 (0.37, 1.4) |
|  | Day 60 | 0.14 (0.05, 0.42) | 0.63 (0.33, 1.23) |
|  | Day 90 | 0.25 (0.1, 0.64) | 0.6 (0.3, 1.17) |
|  | Day 120 | 0.34 (0.17, 0.7) | 0.42 (0.21, 0.84) |
|  | Day 150 | 0.27 (0.13, 0.54) | 0.37 (0.18, 0.75) |
| 65+ years<br>(344 infections,<br>2,821 hospitalizations) | Day 4 | 1 (Reference) | 1 (Reference) |
|  | Days 10 | 0.91 (0.32, 2.62) | 1.06 (0.58, 1.94) |
|  | Day 15 | 0.78 (0.32, 1.91) | 1.12 (0.67, 1.87) |
|  | Day 21<br>(Expected second<br>dose) | 0.68 (0.25, 1.86) | 0.95 (0.57, 1.59) |
|  | Day 35<br>(Expected full<br>vaccination) | 0.21 (0.07, 0.63) | 0.97 (0.58, 1.6) |
|  | Day 60 | 0.14 (0.04, 0.49) | 1.03 (0.61, 1.72) |
|  | Day 90 | 0.05 (0.01, 0.19) | 0.73 (0.43, 1.24) |
|  | Day 120 | 0.22 (0.07, 0.71) | 0.69 (0.4, 1.18) |
|  | Day 150 | 0.18 (0.06, 0.5) | 0.58 (0.34, 1.01) |

**Table S2.** Demographic and clinical characteristics of cases and controls for the sensitivity analysis of BNT162b2 with stratification on county, time of vaccination, and county-level COVID-19 incidence at the time of testing rather than on county and time of testing. The underlying population corresponds to the set of individuals who received their first BNT162b2 dose on or after January 1, 2021 and were fully vaccinated per protocol (i.e. with two doses administered 18-28 days apart and with no prior positive SARS-CoV-2 PCR tests before the date of full vaccination). The eligible population corresponds to the subset of the underlying population who underwent at least one symptomatic test after the date of full vaccination. Cases correspond to the first positive symptomatic test for a given individual in the eligible population; by definition, the number of individuals contributing cases is the same as the number of cases. Controls correspond to negative symptomatic tests after full vaccination which occur before the given individual has experienced any positive SARS-CoV-2 PCR tests; an individual can contribute multiple controls during the study period, so the number of individuals in the control population is less than the total number of tests (controls) contributed. Because an individual can contribute negative tests (controls) prior to contributing a positive test, the number of individuals in the eligible population is smaller than the sum of the number of individuals in the case and control populations. Sub-sampling in the control population refers to the process in which negative tests from a given individual were (i) excluded if they occurred after a positive test or within the 15 days before a positive test (possible false negative), (ii) randomly sampled if they occurred within 15 days of each other (possibly during the same symptomatic illness), and (iii) randomly sampled if the individual contributed more than three negative tests during the study period. A stratum (defined by the regression equation as a unique combination of county and calendar week of testing) is considered analyzable if it includes at least one case and at least one control, because strata including only cases or only controls do not contribute to the estimation of the regression coefficients. For cases and controls, all summarized characteristics correspond to only individuals who contributed at least one symptomatic test to an analyzable stratum.

| Characteristic | Underlying Population | Eligible Population | Case Population | Control Population |
| --- | --- | --- | --- | --- |
|  | Fully vaccinated per-protocol | Fully vaccinated per-protocol with subsequent symptomatic test | 1+ positive symptomatic test after full vaccination | 1+ negative symptomatic test after full vaccination, sub-sampled |
| Number of individuals | 97487 | 17641 | 1008 | 16709 |
| Number of symptomatic tests |  | 21973 |  |  |
| Number of symptomatic tests in analyzable strata |  |  |  |  |
| *Total* |  |  | 974 | 11371 |
| *Bucketed by time since full vaccination date* |  |  |  |  |
| - Days 0-29 |  |  | 46 | 2006 |
| - Days 30-59 |  |  | 58 | 1666 |
| - Days 60-89 |  |  | 150 | 1956 |
| - Days 90-119 |  |  | 263 | 2380 |
| - Days 120-149 |  |  | 204 | 1662 |
| - Days 150+ |  |  | 253 | 1701 |
| Number of individuals contributing symptomatic tests in analyzable strata |  |  | 974 | 10282 |
| Age (years) |  |  |  |  |
| - Mean (sd) | 57.9 (18.7) | 58.6 (19.6) | 58.0 (18.3) | 57.6 (19.8) |
| - 18-24 | 5,243 (5.4%) | 856 (4.9%) | 42 (4.3%) | 485 (4.7%) |
| - 25-34 | 9,675 (9.9%) | 1,984 (11.2%) | 77 (7.9%) | 1,313 (12.8%) |
| - 35-44 | 11,167 (11.5%) | 2,134 (12.1%) | 140 (14.4%) | 1,362 (13.2%) |
| - 45-54 | 12,584 (12.9%) | 2,109 (12.0%) | 153 (15.7%) | 1,253 (12.2%) |
| - 55-64 | 19,513 (20.0%) | 3,076 (17.4%) | 187 (19.2%) | 1,707 (16.6%) |
| - 65-74 | 19,425 (19.9%) | 3,239 (18.4%) | 167 (17.1%) | 1,781 (17.3%) |
| - 75-84 | 15,135 (15.5%) | 2,913 (16.5%) | 155 (15.9%) | 1,626 (15.8%) |
| - 85+ | 4,745 (4.9%) | 1,330 (7.5%) | 53 (5.4%) | 755 (7.3%) |
| State of Primary Residence |  |  |  |  |
| - Arizona | 9,155 (9.4%) | 1,369 (7.8%) | 55 (5.6%) | 964 (9.4%) |
| - Florida | 8,844 (9.1%) | 2,389 (13.5%) | 295 (30.3%) | 1,573 (15.3%) |
| - Iowa | 1,347 (1.4%) | 156 (0.9%) | 8 (0.8%) | 11 (0.1%) |
| - Minnesota | 52,829 (54.2%) | 9,205 (52.2%) | 397 (40.8%) | 5,363 (52.2%) |
| - Wisconsin | 21,229 (21.8%) | 4,304 (24.4%) | 217 (22.3%) | 2,369 (23.0%) |
| - Other | 4,083 (4.2%) | 218 (1.2%) | 2 (0.2%) | 2 (0.0%) |
| Sex |  |  |  |  |
| - Female | 55,356 (56.8%) | 10,684 (60.6%) | 564 (57.9%) | 6,338 (61.6%) |
| - Male | 42,125 (43.2%) | 6,956 (39.4%) | 410 (42.1%) | 3,943 (38.3%) |
| - Unknown | 6 (0.0%) | 1 (0.0%) |  | 1 (0.0%) |
| Race |  |  |  |  |
| - Asian | 2,726 (2.8%) | 493 (2.8%) | 19 (2.0%) | 372 (3.6%) |
| - Black / African American | 2,502 (2.6%) | 501 (2.8%) | 38 (3.9%) | 296 (2.9%) |
| - Native American |  |  |  |  |
| - Native Hawaiian/Pacific Islander | 237 (0.2%) | 48 (0.3%) | 2 (0.2%) | 26 (0.3%) |
|  | 84 (0.1%) | 19 (0.1%) | 1 (0.1%) | 12 (0.1%) |
| - White / Caucasian |  |  |  |  |
| - Other | 88,758 (91.0%) | 16,104 (91.3%) | 886 (91.0%) | 9,294 (90.4%) |
| - Unknown | 2,033 (2.1%) | 349 (2.0%) | 17 (1.7%) | 204 (2.0%) |
|  | 1,147 (1.2%) | 127 (0.7%) | 11 (1.1%) | 78 (0.8%) |
| Ethnicity |  |  |  |  |
| - Hispanic or Latino | 3,104 (3.2%) | 601 (3.4%) | 40 (4.1%) | 344 (3.3%) |
| - Not Hispanic or Latino | 92,055 (94.4%) | 16,739 (94.9%) | 921 (94.6%) | 9,749 (94.8%) |
| - Unknown | 2,328 (2.4%) | 301 (1.7%) | 13 (1.3%) | 189 (1.8%) |
| Elixhauser Comorbidity Index |  |  |  |  |
| - 0 | 43,029 (44.1%) | 7,174 (40.7%) | 434 (44.6%) | 4,231 (41.1%) |
| - 1-4 | 28,990 (29.7%) | 4,719 (26.8%) | 292 (30.0%) | 2,861 (27.8%) |
| - 5-9 | 18,164 (18.6%) | 3,580 (20.3%) | 164 (16.8%) | 1,966 (19.1%) |
| - 10+ | 7,304 (7.5%) | 2,168 (12.3%) | 84 (8.6%) | 1,224 (11.9%) |
| Dates of full vaccination |  |  |  |  |
| - Earliest | Feb 3 | Feb 5 | Feb 6 | Feb 5 |
| - 25th %<br>- Median<br>- 75th %<br>- Latest | Mar 5<br>Apr 6<br>May 3<br>Aug 22 | Mar 3<br>Mar 30<br>Apr 25<br>Aug 20 | Mar 3<br>Apr 1<br>Apr 26<br>Jul 22 | Feb 27<br>Mar 18<br>Apr 19<br>Jul 25 |
| Time between full vaccination<br>and test<br>- Minimum<br>- 25th %<br>- Median<br>- 75th %<br>- Maximum |  | 0<br>41<br>83<br>121<br>196 | 0<br>88<br>116<br>152<br>196 | 0<br>45<br>90<br>128<br>196 |

**Table S3.**
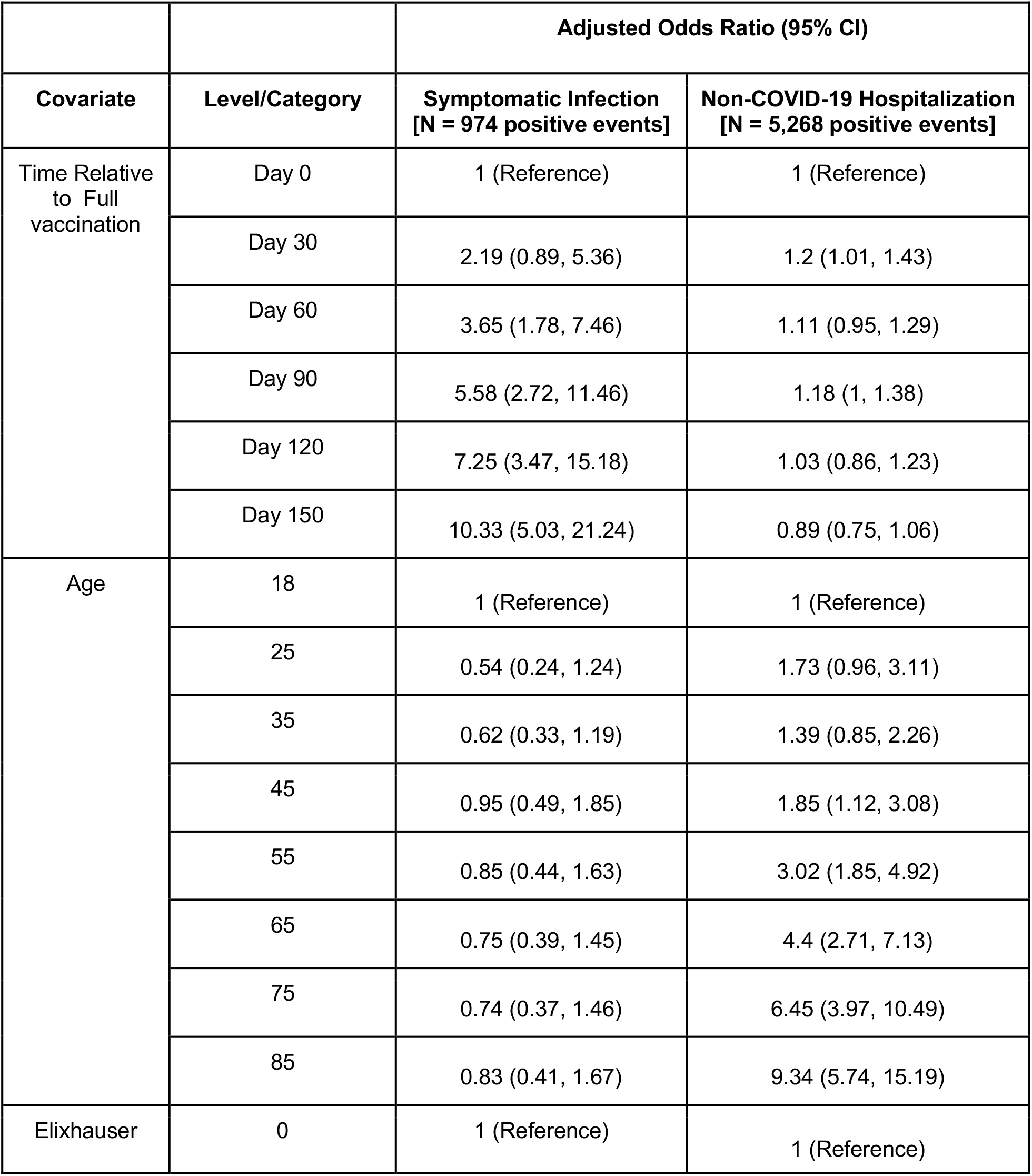
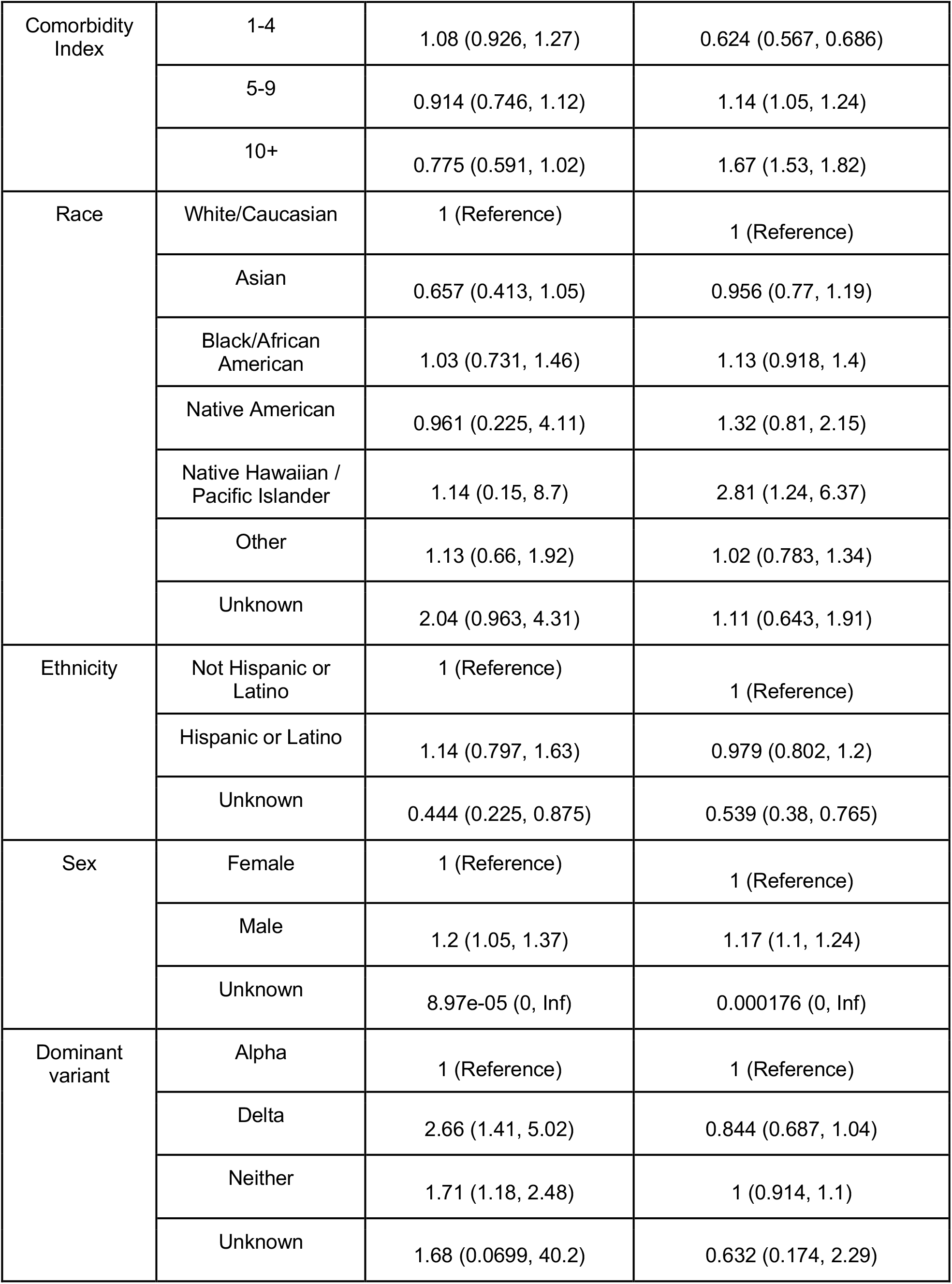
Sensitivity analysis 2: conditional logistic regression with stratification on county, time of vaccination, and county-level COVID-19 incidence at the time of testing rather than on county and time of testing. There were 974 positive symptomatic tests and 5,268 non-COVID-19 hospitalizations that contributed to analyzable strata after the date of full vaccination. Adjusted odds were estimated with linear spline equations (for time since full vaccination and age) or by exponentiating the coefficients derived from conditional logistic regression models fit separately for each outcome. For “Time Relative to Full Vaccination”, knots through the 90th percentile of the days since full vaccination for the entire cohort are included. Related to **Figure S3**.

